# Impact on all-cause mortality of a case prediction and prevention intervention designed to reduce secondary care utilisation: findings from a randomised controlled trial

**DOI:** 10.1101/2022.10.14.22281029

**Authors:** Lucy Bull, Bartlomiej Arendarczyk, An Nguyen, Joachim Werr, Thomas Lovegrove-Bacon, Mark Stone, Chris Sherlaw-Johnson

**Affiliations:** Health Navigator Ltd, The Brew Eagle House, 163 City Rd, London, EC1V 1NR; Health Navigator Ltd, London UK; Health Navigator Ltd, London, UK; East Kent Hospitals University NHS Foundation Trust, Kent, UK; Stafford Health and Wellbeing, Stafford, UK; Nuffield Trust, London, UK

## Abstract

**Objective:** To investigate how an AI case-finding and clinical coaching intervention impacted mortality and how this impact varied by age, gender, and deprivation status.

**Design:** Multi-site parallel prospective two-arm Randomised Controlled Trial led by Nuffield Trust and delivered by HN (Health Navigator Ltd). Patients were randomised on a 2:1 ratio to the intervention after consent and the automated and manual screening processes.

**Setting:** Secondary care-based patient identification for a community-based intervention; Eight hospital sites across England were enrolled onto the study (York, Staffordshire, Essex, and Kent).

**Participants:** Subjects aged 18 and over, who have experienced at least one emergency attendance in the preceding six months and identified as high-risk of unplanned hospitalisation via a prediction model. Subjects were also manually screened for their suitability to intervention.

**Intervention:** One-to-one telephone-based health coaching, led by registered nurses or paramedics.

**Primary outcome measure:** 24-month mortality.

**Results:** The intervention was associated with reduced overall mortality (posterior probability: 92.2%), predominantly driven by the impact for males aged 75 and over (log-rank p-value: 0.0011, Hazard Ratio (HR) [95% CI]: 0.57 [0.37, 0.84], number needed to treat: 8). Excluding one site unable to adopt the prediction model indicated stronger impact (HR [95% CI]: 0.45 [0.26, 0.76]), suggesting a role of prediction in reducing mortality.

**Conclusions:** Early mortality, specifically in elderly males, may be prevented by predicting individuals at risk of unplanned hospitalisation and supporting them with a clear outreach, out-of-hospital nurse-led, telephone-based coaching and care model.

**Trial registration:** IRAS project ID: 173319; and clinicaltrials.gov ID: 2015–000810-23

**Key messages:** *What is already known on this topic:*

- The overcrowding of emergency departments is a major global issue that has motivated the development of alternative models of care (e.g., case management interventions) to both reduce the strain on hospitals and improve health outcomes.
- Existing interventions, designed to reduced unplanned secondary care and improve patient outcomes, are rarely evaluated for their impact on mortality.

*What this study adds:*

- A large parallel multi-site randomised controlled trial, involving 1688 patients, suggested that an AI case-finding and clinical coaching intervention, can reduce mortality rates for males aged 75 and over.
- Excluding one site technically unable to adopt the prediction model provided stronger impact, suggesting a role of prediction in reducing mortality.

*How this study might affect research, practice, or policy:*

- Predicting unplanned hospitalisation using routinely collected secondary care data, and supporting at-risk patients earlier with remote, anticipatory care could help save lives, and address gender-related health inequalities.

## 1. BACKGROUND

The overcrowding of emergency departments is a major global issue, which has been further exacerbated by the lack of in-person services during the COVID-19 pandemic [1], having a detrimental effect on health outcomes by prolonging access to relevant care [2].

The causes of emergency department crowding are complex and multifactorial [3]. However, it is widely known that many emergency visits are avoidable, with 24% of them being turned away without treatment or with advice only in the UK [4, 5]. Furthermore, a small number of patients account for a disproportionately large amount of non-elective secondary care [6], with the top 5% frequent users accounting for more than 25% of emergency department visits [7].

Promising evidence, to reduce demand of frequent users on emergency services, is emerging for community-based services and interventions that improve access to primary care services and promote the continuity of care [8-10]. A wide range of interventions to achieve these outcomes have been proposed (e.g., from health coaching to virtual monitoring [11-13]), but the quality of evidence varies [14]. Frequent users are also highly transient over time, with individual patients typically returning to the expected (or lower) secondary care demand within 12 months [15]. Thus, impact evaluations require a matching control group, and an effective data-driven approach is required to identify the amenable target population in advance of a clinical crisis [16, 17].

Although existing interventions are designed and evaluated to reduce healthcare demand and improve patient outcomes [11-14], they are rarely evaluated for their impact on mortality. In the UK, a single study investigated the impact of a virtual monitoring intervention on mortality [13], but limited evidence exists for other interventions [11, 12, 14, 17]. Globally, mortality impact was evaluated for a telephone-based intervention in older adults in managed care [18], for patients with long-term conditions [19-21], and for patients with healthcare utilisation in the previous six months [22, 23].

### 1.2. Aim and objectives

The primary objective of this study was to investigate how an AI case-finding and clinical coaching intervention, prioritised for patients identified as high risk of a clinical crisis and unplanned hospitalisation by a prediction model, impacted mortality. The secondary objective was to provide a more granular insight into how the mortality impact varied by age, gender, and deprivation status.

## 2. METHODS

All methods and results have been reported in line with the CONSORT reporting checklist, see Appendix D.

### 2.1. Data

#### 2.1.1. Setting

This was a multi-site parallel two-arm randomised control trial (RCT) where Nuffield Trust was the principal investigator, and the intervention was conducted by Health Navigator Ltd (Integrated Research Application System project ID: 173319; and clinicaltrials.gov ID: 2015–000810-23). The study was designed to formally evaluate the impact of a telephone-based case-management health coaching intervention using an algorithm-generated selection of patients at elevated risk of future unplanned secondary care. Recruitment started in August 2015 and concluded in 2019.

#### 2.1.2. Study population and randomisation

Patient-level data was obtained from each study site (York Teaching Hospitals, East Kent Hospitals University Foundation Trust, University Hospitals Derby and Burton, University Hospitals North Midlands, Royal Wolverhampton Trust, Mid and South Essex NHS Foundation Trust) and linked with the NHS Spine dataset to obtain both inpatient and outpatient reported deaths. NHS Spine data, managed by NHS Digital, refers to a linked collection of local and national databases and systems containing both a patient’s clinical and demographic information [24]. Included patients were those at high risk of 3 unplanned bed days or more (see Appendix B.2), at least 18 years old and had an emergency attendance in the preceding six months. Patients were manually screened for their suitability to intervention, relating to their medical diagnoses and ability to benefit from a telephone-based service (see Appendix B.1.1). Included patients were invited to participate in the trial. Consented patients were openly randomised using an online random-sequence generator to either the intervention or control group using a 2:1 ratio in favour of the intervention. Recruited patients were observed for 24 months post-randomisation.

#### 2.1.3 Sample size

The RCT study was initially powered to detect a clinically meaningful 12-15% reduction in emergency admission rates with 90% statistical power (see Appendix B.1.3). Although the initial power calculations suggested an overall sample of 3000, local principal investigators and the Nuffield Trust mutually agreed to close recruitment due to decreasing recruitment rates and service providers no longer supporting the withholding of the intervention from control patients.

#### 2.1.4. Intervention

Patients in the intervention group received an initial face-to-face assessment meeting with a Health Navigator health coach, who are registered nurses or paramedics, to better understand their health and social care needs and establish any gaps in the care provided by the system. The initial assessment promoted the development of a personalised care plan and was followed by a telephone-based case management intervention, which comprised of one-to-one telephone calls with the coach. During these calls, patients received motivational guidance, support for self-care, health education, and coordination of social and medical services. Motivational guidance, informed by existing theories (e.g.,[25]); components included demonstrating empathy, dealing with resistance, supporting self-efficacy, and developing autonomy. No medical advice or treatment was delivered. Patients in the control group received NHS standard care, defined here as the primary and community services already in place within the area.

#### 2.1.5. Data collection and management

The RCT study participants were linked to the NHS Spine data using their NHS number. Patients were excluded from the analysis if they were unable to be linked to the Spine data, or if they had missing age, sex, or deprivation status. A follow-up observation period of two-years from the point of randomisation was included for the evaluation of mortality status.

### 2.2. Statistical analysis

This section reports key statistical methods employed for the primary and secondary analyses. A more detailed overview can be found in Appendix C. Analysis was performed in R version 4.2.1.

#### 2.2.1. Descriptive analysis

To informally compare patient demographic case-mix across treatment groups identified via randomisation, summary statistics were provided for age, sex, and Index of Multiple Deprivation (IMD; as a measure of social economic status) decile. Crude rates were compared across groups Kaplan-Meier curves. Kaplan-Meier curves were provided for patients in the intervention and control groups, and for age, gender and IMD sub-groups. Age was divided into two categories for pairwise interactions, under the age of 75 and aged 75 and over. The IMD deciles were divided into two categories, the 50% most deprived areas (IMD decile less than or equal to 5) and 50% least deprived areas (decile > 5).

#### 2.2.2. Statistical modelling

A Bayesian Weibull survival model [26] was fitted with fixed effects for the intervention group (binary indicator), the top 50% most deprived (binary indicator), age (in decades, and above/below 75 years) and sex, and a random effect (frailty) for the intervention site. Although age was divided into categories when investigating pairwise interactions to aid interpretation, it was included as a continuous variable when employed as a control. The random effect accounts for variation in the baseline hazard function between the different intervention sites and assumes proportionality between them. Non-informative priors were assumed for all the fixed effects except for the intervention indicator and interactions. Prior studies evaluating similar interventions suggested little impact on mortality. Thus, for the group indicator, an informative prior assuming a null effect of the intervention on mortality was imposed with a variability that ensured that approximately 95% of the density function lay between a hazard ratio of 0.2 and 5. The derivation of the informative prior is provided in Appendix C.1.1.1. First-order two-way interactions between the intervention indicator and each of the control variables were added into the model, and then second-order interactions between age and gender were investigated given the first-order interactions.

#### 2.2.3. Numbers needed to treat

The two-year Absolute Risk Reduction (ARR) and associated Number Needed to Treat (NNT) to prevent one death were calculated to provide a clinically interpretable measure of the impact of the intervention on mortality.

### 2.3. Sensitivity analysis

To investigate the potential impact of the choice of prior and the parametric survival function, the primary analysis was performed again with a non-informative prior for the intervention group and using a Cox proportional hazards model. In addition, as one of the sites did not have the technical readiness required for prediction model implementation, the primary analysis was performed again without the data from this site. In 2017, the digitalisation of paper records at Mid Essex Hospital Services NHS Trust had only just started. Consequently, limited historical patient information could electronically feed into the model.

To explore potential explanations to the findings of the primary analysis, secondary analysis was performed on intervention subjects using the data collected by the clinical coaches, and on primary care utilisation data on a subset of the York participants. To gain insight into the social background of the intervention subjects, proportions of males aged 75 and over living alone were compared to rest of the intervention arm. To gain insight into the medical profiles of intervention subjects, patient-reported clinical diagnoses were compared for males aged 75 and over and the rest of the intervention arm. Comparisons could not be made to the control group due to how the data was generated. To explore whether the intervention led to an increase in primary care utilisation, incidence rates were compared across subgroups of interest.

## 3. RESULTS

The CONSORT flow diagram to illustrate the inclusion process for the study population is presented in Figure 1.

**Figure 1:**
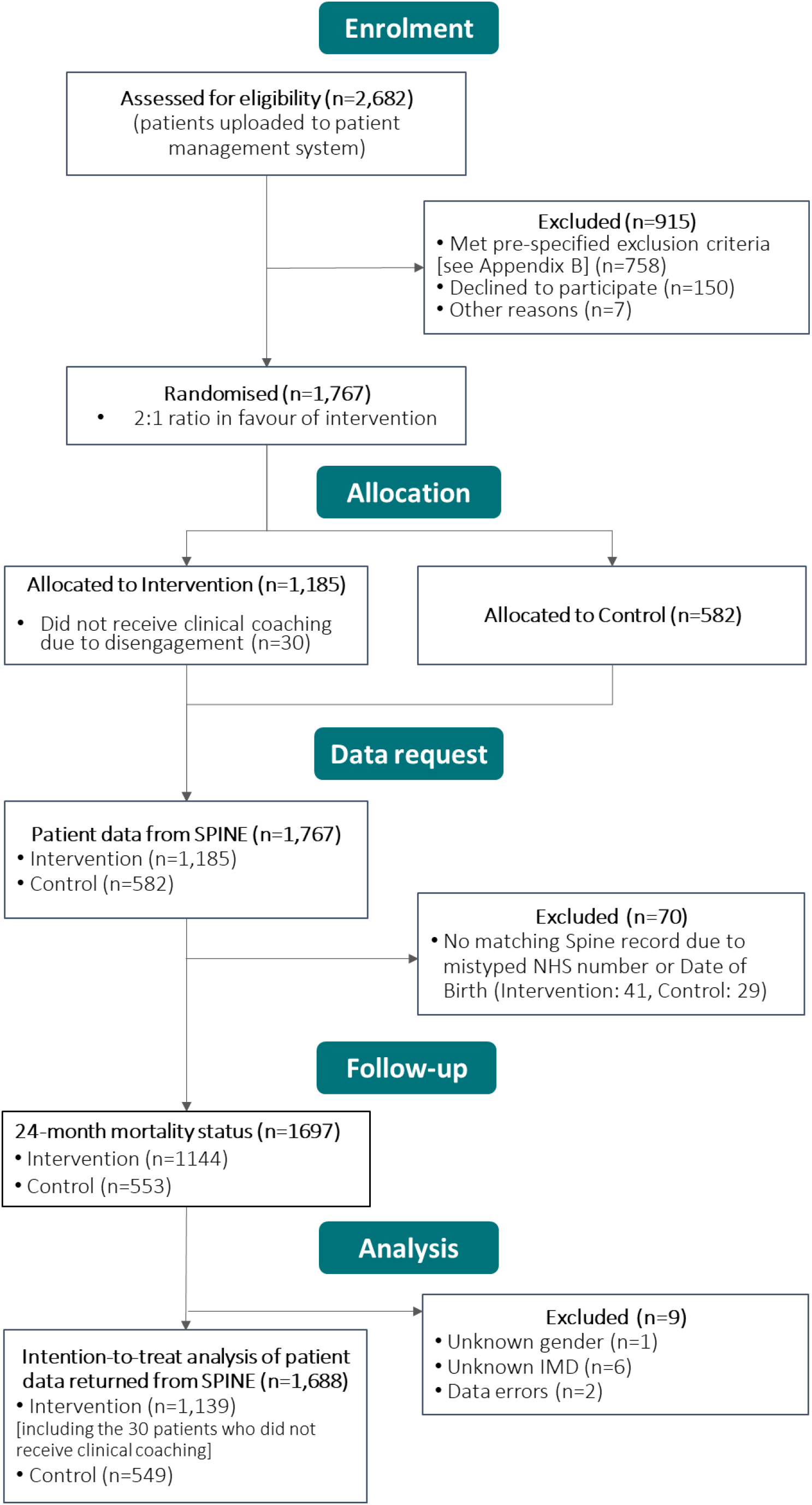
CONSORT flow diagram to illustrate the inclusion process for the study population.

### 3.1. Descriptive analysis

The baseline characteristics of the study participants are presented in Table 1. These findings suggest that there was a similar distribution of age categories, gender, and deprivation status across both treatment arms. Both groups have a median age of 75 (interquartile range (IQR) = 15 and 14), an even balance of genders (52.8% vs 52.5% female), and just under a third of the group living in the top 50% most deprived areas (30.8% vs 29.3%).

**Table 1:** Summary statistics of age, gender, and deprivation status for the overall study population and for the intervention and control groups, as well as numbers needed to treat (NNT) for sub-groups of interest [IQR – Interquartile range].

|  | Intervention<br>(n= 1139) | Control<br>(n= 549) |  |  |
| --- | --- | --- | --- | --- |
| Age, median (IQR) | 75 (15) | 75 (14) |  |  |
| Age categories, count (%) |  |  |  |  |
| Under 75 | 546 (47.9) | 258 (47.0) |  |  |
| 75 and over | 593 (52.1) | 291 (53.0) |  |  |
| Female, count (%) | 601 (52.8) | 288 (52.5) |  |  |
| Lives in 50% most<br>deprived area, count<br>(%) | 351 (30.8) | 161 (29.3) |  |  |
| Sites, count (%) |  |  |  |  |
| Burton | 34 (3.0) | 17 (3.1) |  |  |
| East Kent | 90 (7.9) | 54 (9.8) |  |  |
| Mid-Essex | 267 (23.4) | 137 (25.0) |  |  |
| North Midlands | 79 (6.9) | 33 (6.0) |  |  |
| Wolverhampton | 117 (10.3) | 57 (10.4) |  |  |
| York 1 | 163 (14.3) | 60 (10.9) |  |  |
| York 2 | 389 (34.2) | 191 (34.8) |  |  |
| Mortality (deaths/1000 patients-years) (N) |  |  | Difference<br>(%) | NNT |
| Overall | 64.7 (1139) | 80.1 (549) | -19.2 | 38 |
| Gender |  |  |  |  |
| Female | 54.5 (601) | 53.0 (288) | 2.8 | * |
| Male | 76.3 (538) | 112.3 (261) | -32.1 | 17 |
| Age |  |  |  |  |
| Under 75 | 46.1 (546) | 42.6 (258) | 8.2 | * |
| 75 and over | 82.5 (593) | 115.9 (291) | -28.8 | 18 |
| Deprivation |  |  |  |  |
| 50% most deprived | 72.0 (351) | 84.4 (161) | -14.7 | 47 |
| 50% least deprived | 61.5 (788) | 78.4 (388) | -21.6 | 35 |
| Age & Gender |  |  |  |  |
| Female, Under 75 | 27.6 (298) | 33.3 (140) | -17.1 | 94 |
| Female, 75 and over | 82.6 (303) | 72.2 (148) | 14.4 | * |
| Male, Under 75 | 69.3 (248) | 53.9 (118) | 28.6 | * |
| Male, 75 and over | 82.4 (290) | 166.4 (143) | -50.5 | 8 |
|  |  | * no reduction in mortality |  |  |

Comparing crude mortality rates using Kaplan-Meier curves suggested that the largest reduction in mortality across treatment arms was for males aged 75 and over (log-rank p-value: 0.0011). The difference in rates was statistically significant according to the log-rank test (see Figure 2). Kaplan Meier curves for overall treatment arms and other subgroups of interest can be found in Appendix C; these suggested an overall reduction in mortality for the intervention group (log-rank p-value: 0.13), for male patients based in the 50% least deprived areas (log-rank p-value = 0.050), and for patients 75 and over in the 50% least deprived areas (log-rank p-value = 0.037).

**Figure 2:**
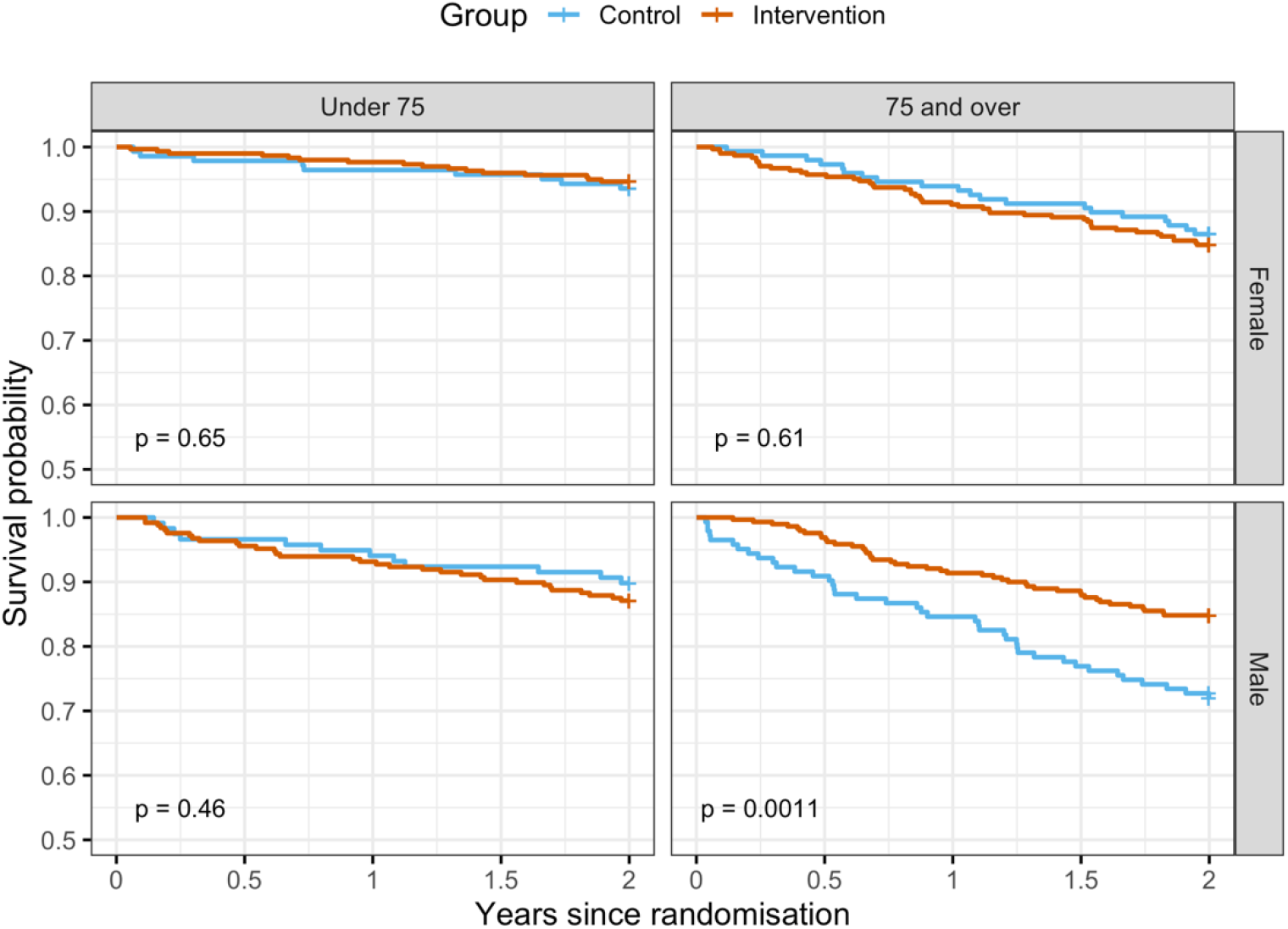
**Kaplan-Meier curves the intervention (orange) and control (blue) groups across the different age (across the top) and gender (down the right-hand side) bands included in the models**.

### 3.2. Statistical summary and modelling

Estimated hazard ratios (HR) and credible intervals (CI) relevant to the Bayesian Weibull survival models (with an informative prior) can be found in Table 2. The posterior distributions of the estimated hazard ratios are illustrated in Figure 3, this includes those for the first order (pairwise) and second order (three-way) interactions of interest.

**Table 2:** An overview of findings using the Weibull survival model for both overall mortality and sub-group analyses using an informative prior.

|  | Hazard Ratio [95% Credible Interval] | Posterior<br>P(HR < 1) |
| --- | --- | --- |
| **Model 1 – Overall** |  |  |
| Intervention | 0.82 [0.62, 1.08] | 0.92 |
| Age (decades) | 1.60 [1.38, 1.89] | - |
| Gender=M | 1.66 [1.26, 2.23] | - |
| Deprivation=High | 1.24 [0.92, 1.67] | - |
| **Model 2 - By gender** |  |  |
| Intervention:Male | 0.71 [0.50, 1.01] | 0.97 |
| Intervention:Female | 1.02 [0.67, 1.62] | 0.46 |
| Age (decades) | 1.60 [1.38, 1.90] | - |
| Gender=M | 2.10 [1.34, 3.44] | - |
| Deprivation=High | 1.25 [0.92, 1.69] | - |
| **Model 3 - By age categories** |  |  |
| Intervention:Age[0,75) | 1.06 [0.70, 1.61] | 0.39 |
| Intervention:Age[75,100) | 0.72 [0.52, 0.99] | 0.98 |
| Age (decades) | 1.77 [1.46, 2.19] | - |
| Gender=M | 1.66 [1.27, 2.24] | - |
| Deprivation=High | 1.24 [0.91, 1.67] | - |
| **Model 4 - By deprivation bands** |  |  |
| Intervention:HighDeprivation | 0.90 [0.55, 1.48] | 0.67 |
| Intervention:LowDeprivation | 0.79 [0.56, 1.10] | 0.91 |
| Age (decades) | 1.60 [1.38, 1.90] | - |
| Gender=M | 1.66 [1.26, 2.23] | - |
| Deprivation=High | 1.14 [0.69, 1.83] | - |
| **Model 4 - By age and gender** |  |  |
| Intervention:Age[0,75):Female | 0.97 [0.52, 1.78] | 0.54 |
| Intervention:Age[75,100):Female | 1.03 [0.65, 1.65] | 0.45 |
| Intervention:Age[0,75):Male | 1.10 [0.68, 1.78] | 0.34 |
| Intervention:Age[75,100):Male | 0.57 [0.37, 0.84] | 1.00 |
| Age (decades) | 1.76 [1.45, 2.19] | - |
| Gender=M | 2.10 [1.36, 3.37] | - |
| Deprivation=High | 1.25 [0.92, 1.70] | - |

**Figure 3:**
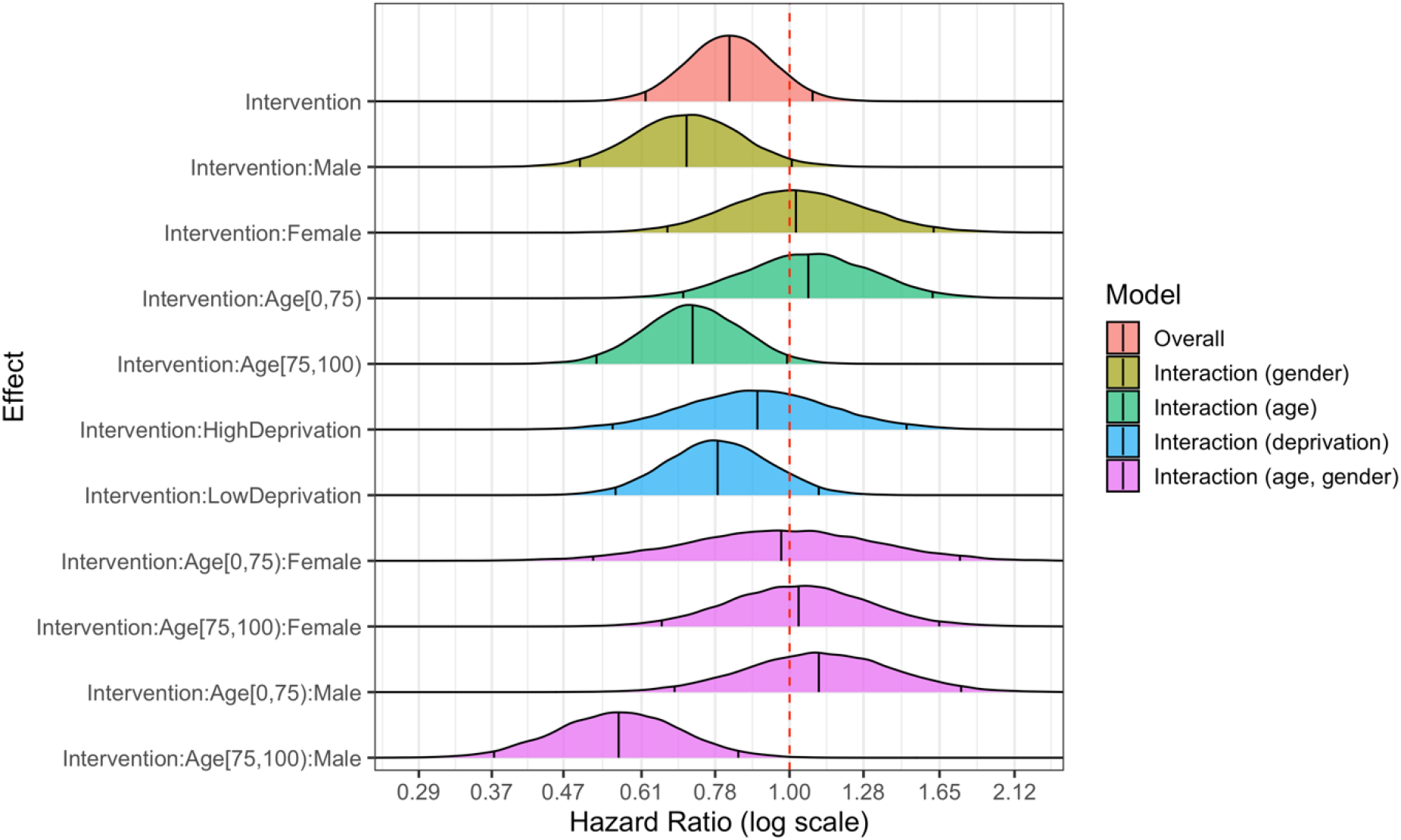
**Illustration of the posterior distributions for the hazard ratios for models exploring overall mortality impact of the intervention (red), pairwise interactions with gender (yellow), age (green) and deprivation band (blue), and second-order interactions with gender and age (violet)**.

According to the results presented in Table 2, the intervention group showed lower overall mortality compared to the control group (HR=0.82, 95% CI=[0.62, 1.08]). Male patients showed a strong reduction in mortality (HR=0.71, 95% CI=[0.50, 1.01]) whereas females showed little difference in mortality compared to the control (HR=1.02, 95% CI=[0.67, 1.62]). Among the two age categories, a reduction in mortality was only observed in patients aged over 75 years (HR=0.72, 95% CI=[0.52, 0.99]). The strongest reduction in mortality was observed in male patients, aged 75 and over (HR=0.57, 95% CI=[0.37, 0.84]), thus suggesting that the reduction in mortality rate indicated by the pairwise interactions can be explained by this sub-group of patients. A stronger association with reduced mortality was identified for patients receiving the intervention in the 50% least deprived areas (HR: 0.79, CI= [0.56, 1.10]) compared to those in the 50% most deprived (HR: 0.90, CI= [0.55, 1.48]), but a wide credible interval for those in the highly deprived areas suggests prominent levels of uncertainty and smaller numbers in this subgroup.

#### 3.2.1. Sensitivity analysis

The sensitivity analyses performed suggested consistent findings across the choice of prior, and statistical modelling approach (crude mortality reduction of 50.5% for males aged 75 and over, non-informative prior HR: 0.54 [0.35, 0.83] and Cox PH HR: 0.55 [0.37, 0.83], see Table C.3 in Appendix C for all findings). However, stronger reductions in overall mortality rates for males aged 75 and over in the intervention group were observed once the Mid-Essex site participants were removed from the analysis (HR: 0.45 CI= [0.26, 0.76] without Mid-Essex compared to HR:0.57 CI= [0.37, 0.84]), see Table C.4 in Appendix C). The secondary analysis on intervention subjects suggested that the mortality impact observed on males aged 75 and over may not be explained by their living situation or their medical profile but comparisons to matching controls are required (see Appendix C.4.2.1). An increased primary care consumption was also not observed for this sub-group of interest (see Appendix C.4.2.2).

## 4. DISCUSSION

### 4.1. Overall summary of findings

For patients identified by a predictive algorithm to be at elevated risk of increased hospital utilisation, the findings suggested that the intervention was associated with an overall mortality reduction (posterior probability of 92.2%), predominantly for males aged 75 years and over. Stronger associations with reduced mortality were estimated when a single site, unable to employ the prediction model, was excluded suggesting that prediction could play a key role in mortality impact.

### 4.4 Comparison with existing literature

Despite the association with reduced mortality being unexpected, the findings of this study align with previous evidence for health coaching interventions for those with multiple chronic conditions [22], cardiovascular disease [21], and for elderly patients in managed care [19]. Null effects on mortality were found in other studies, however [20, 23, 24]. Granular investigations of the association between the intervention, survival and sub-groups of the study population are extremely limited with most studies focussed on the overall impact on patient mortality [19, 20, 22]. A single study reported a greater association between the intervention and reduced mortality for those with more coaching sessions, and for those who were male, but the intervention was evaluated on patients diagnosed with cardiovascular disease only [21].

It is difficult to understand the impact of data-driven approaches in previous literature, as they have not been combined with health coaching in the same way. Mortality evidence of a rule-based approach with a collaborative primary-care based intervention suggested reduced mortality [27].

### 4.2. Study Limitations

It is important to acknowledge the study limitations when interpreting the findings. First, as the study population were identified by a predictive algorithm, they are likely to be sicker and more vulnerable than the general population. This would not be an issue for generalisability, however, as the algorithm to identify suitable subjects would be combined with the clinical coaching intervention. Second, access to further data on the study population was limited due to accessibility issues from some RCT study sites. Using this data, it would have been beneficial to control for primary and secondary care utilisation, but data linkage issues prevented this information being incorporated in the Bayesian models. To highlight the bias introduced by these sub-populations, sensitivity analyses were performed. The lack of primary care linkage also prevented a formal evaluation of case-mix consistency across the intervention and control groups. However, any differences between the groups were determined by chance and highly unlikely given the extended screening process and randomisation. Third, the study was not initially powered for mortality as it was not a primary outcome for the intervention. Nevertheless, strong associations were identified in a subset of the study population and Bayesian inference was employed to aid with clinical interpretability of the findings given the available data. Fourth, some patients were excluded from the study due to missing demographics or linkage problems with the Spine dataset, but these numbers were small.

### 4.5 Impact and implications

This study has shown that a prediction and prevention intervention, designed to reduce secondary care utilisation, is associated with a positive impact on survival rates for those who are identified as likely to increase their healthcare consumption, male and aged 75 and over. It is important to acknowledge that the study findings do not suggest that the intervention should only be implemented for those who are male and aged 75 and over, as secondary care utilisation and patient-reported outcomes are the primary endpoints of evaluation.

The sensitivity analyses performed suggested that a similar proportion of elderly males were living alone compared the rest of the intervention group, elderly males did not consume more primary care than controls or the rest of the intervention group, nor did they present with different patient-reported medical conditions with the clinical coaches compared with the rest of the intervention arm. Thus, given our investigations, the potential biases and existing evidence in this area, the authors believe that the mortality impact for elderly males could be due to the intervention (1) identifying potentially vulnerable patients in a timely manner using data-driven approaches [28], (2) being more proactive in enabling access to health services for those more vulnerable due to health inequalities [29], and (3) ‘recruiting’ patients for extra support rather than ‘referring’ them onto other services [30]. The findings of this study highlight health inequalities, driven by patient gender, which receive minimal attention in the literature [29].

Further investigations using the full RCT dataset, once accessible, are required to establish the core elements of the intervention that are driving the mortality impact, but overall, this study currently suggests that it has the potential to save lives.

## 5. CONCLUSION

This is the first UK study demonstrating a significant survival impact of a data-driven prediction and prevention model for elderly patients. This study has shown that an intervention, designed to reduce secondary care utilisation, can have a positive impact on survival rates for those who are identified as likely to increase their healthcare consumption, male and aged 75 and over. Thus, mortality, specifically in elderly males, could be prevented by predicting individuals at risk of unplanned hospitalisation and supporting them with a clear outreach, out-of-hospital care model.

## Data Availability

All data requests should be directed to Health Navigator via

## ACKNOWLEDGEMENTS

The authors would like to note the outstanding efforts of the health coaching nurses and other staff involved in the provision and development of this intervention who made this study possible, and of course the patients. Special thanks also go to the principal investigators at the each of the study sites for their substantial contribution to the delivery and success of the trial (In particular, for York; Dr Nigel Wells, George Scott, and Fiona Bell Morritt. For East Kent: Dr Marc Farr. For Staffordshire Dr Paddy Hannigan). The authors would like to thank Jamie Davis for the natural language processing analysis performed to exchange free text data to SNOMED codes as reported in the supplementary material. The authors would also like to thank Graham Prestwich, Patient involvement and engagement lead at Yorkshire and Humber Academic Health Science Network for leading and delivering the patient involvement to supplement the presented analysis.

## DATA SHARING STATEMENT

All data requests should be directed to Health Navigator via

## PATIENT AND PUBLIC INVOLVEMENT

Patient perceptions of the intervention evaluated as part of this RCT were published online elsewhere to better understand patient concerns and ensure the service was reaching patient need (https://d13bz8i7p8siyq.cloudfront.net/uploads/downloads/Public-and-patient-feedback-on-the-benefits-of-AI-guided-clinical-coaching-Report-Final.pdf).

## ETHICS STATEMENTS

### Patient consent for publication

All patients recruited onto the trial consented to the publication of findings that emerge from the patient-level data.

### Ethics approval

The study has received ethical approval from the NHS Research Ethics Committee and is registered with the National Institute for Health Research (Central Portfolio Management System reference: 19857) and Health Research Authority (Integrated Research Application System ID: 173319).

## AUTHOR CONTRIBUTIONS

LB, BA, AN, TLB and JW conceived the study. LB and BA developed the study design and analytical approach, in consultation with other project team members. TLB performed initial analyses and extracted the relevant data sources. BA performed the statistical analyses, which were supervised by LB, AN, and JW. Data and coding scripts were quality assured by CSJ. LB drafted the initial version of the manuscript. All authors contributed to the interpretation of the findings and reviewed and edited the manuscript for intellectual content. All authors approved the final version of the manuscript and agreed to be accountable for all aspects of the work. The corresponding author attests that all listed authors meet authorship criteria and that no others meeting the criteria have been omitted.

