## Appendix for "Impact on all-cause mortality of a case prediction and prevention intervention designed to reduce secondary care utilisation: findings from a randomised controlled trial"

### Supplementary information

#### Contents

*Page*

**Appendix A – Layman summary**……………………………………..………2
**Appendix B– Study and intervention design**
B.1 Randomised Controlled Trial design……………………………………………….… 3
B.1.1 Manual screening of high-risk patients……………..……………………… 4
B.1.1.1. Inclusion and exclusion criteria……………………………………. 4
B.1.2 Randomisation……..……………………………………………………….. 4
B.1.3 Sample size for RCT primary endpoint……………………..……………… 4
B.1.4 Patient recruitment and consent……………………..…….….………….. 5
B.1.5 Intervention..………….…………………………………………….……… 5
B.2 Prediction model development and validation overview………………………….……… 5

**Appendix C– Supplementary statistical methods and findings**
C.1 Descriptive analysis………………………………………………………………… 7
C.1.2 Methods…………………………………………………………………7
C.1.2 Findings ……………………..……………………………………………... 7
C.2. Primary analysis……………………………………………………………..……….. 8
C.2.1 Methods…………………………………………………………..…… 8
C.4.1.1 Patient-reported living situation and medical conditions ………..… 16
C.4.1.2 Primary care consumption for a subset of York RCT participants… 16
C.4.2 Findings ……………………………………………………………..…. 16
C.4.2.1 Patient-reported living situation and medical conditions ……..…… 16
C.4.2.2 Primary care consumption for a subset of York RCT participants… 20

**Appendix D: CONSORT Reporting Checklist**…………………..……………… .23

**References** ........................................................................................................... 25

#### *Appendix A – Layman summary*

The overcrowding of emergency departments is a huge problem around the world, which was worsened by the COVID-19 pandemic. For this reason, researchers are constantly seeking new ways to help patients outside of hospital to prevent long waiting times in emergency departments. Extended hospital waiting times can prevent patients from getting the care they need on time. Services that can help vulnerable patients, who tend to regularly attend emergency departments for non-emergency related problems, outside of the hospital setting can reduce the amount these patients visit hospital unnecessarily. However, it is currently unknown whether such services could also help save lives.

In this study, we offered a health coaching service to patients who were identified (from their hospital records) as likely to attend hospital unexpectedly soon after. The service involved one-to-one telephone calls with a coach to help address any problems the patients were having with getting the care they needed. Although the coaches did not provide medical advice, they offered support in communicating with health services outside of the hospital, and motivational guidance to help the patients access all the tools they need to confidently manage their own health.

We had two groups of patients from across England, those who were received the service and those who did not, but all patients were suitable for it. When comparing the number of patients who were still alive at the end of the two years between the two groups, we found that more males aged 75 and over survived who received the service compared to those that did not. Although the difference that was observed in this study could be explained by a range of different biases, the authors did their best to explain them using the patient’s living situation, their reported medical profiles, and the number of times they visited their GP.

If the findings of this study do hold, they suggest that combining artificial intelligence to find suitable patients and clinical coaching in an intervention could help save the lives of elderly males who are unlikely to be mitigating risk or accessing the care they need. The number of males aged 75 and over and at elevated risk of a clinical crisis needed to treat with this service is only 8 to save a single life according to the findings in this study. This number can, inevitably, vary from city to city, however.

If we extrapolate these findings to the UK population, there were 2,490,900 males aged 75 and over in 2020 (ONS). Around 3-5% of this group would have been defined as moderately to severely frail by Age UK (2019) and could be eligible to receive the service. With an estimated decrease in mortality between 16% and 63% associated with the service from the study, between 4,184 and 22,678 lives could be saved within two years of enrolment into the service (for every 74,727 to 124,545 enrolled).

### *Appendix B – Study and intervention design*

#### B.1. Randomised Controlled Trial design

An overview of the data-driven patient recruitment and clinical coaching intervention can be found in Figure B.1. A more detailed overview of the inclusion process and how it aligns with the process illustrated in Figure B.1 can be found in Figure B.2. The remainder of this sub-section details the manual screening process, as well as the randomisation and consent processes. The subsection concludes with more details about the telephone-based coaching intervention itself.

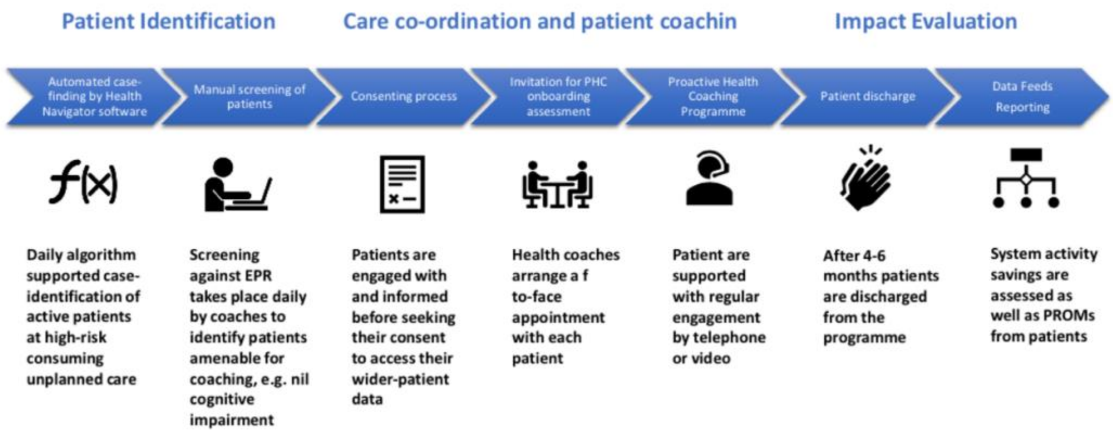

**Figure B.1 An overview of the data-driven patient recruitment and case-management intervention**

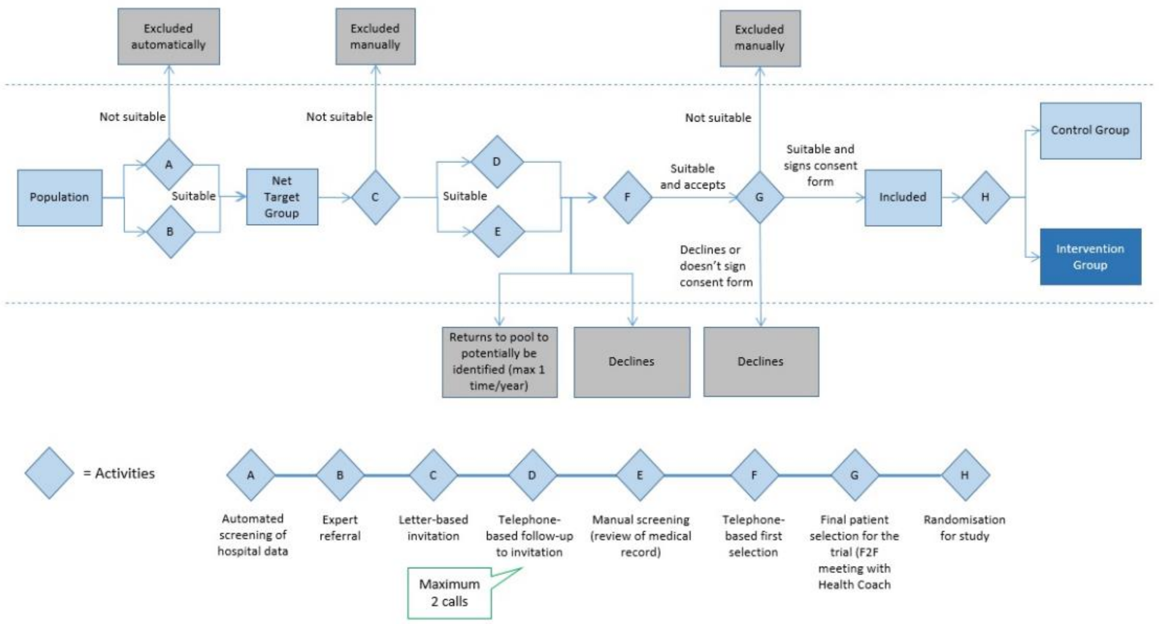

**Figure B.2 A detailed overview of the inclusion process and how it aligns with the RCT recruitment process**

##### B.1.1 Manual screening of high-risk patients

After the initial identification of patients via the prediction model (described later in Section B.2), the identified subjects were then manually screened for their suitability to the service provided. The HN coaches perform the manual screening of the patients using the inclusion and exclusion criteria stated in Section B.1.1.1.

###### B.1.1.1 Inclusion and exclusion criteria

Inclusion criteria:

- Aged 18 or over
- One or more emergency attendances in the preceding six months
- Identified as high-risk of becoming a frequent user by the prediction model

Exclusion criteria:

- Patients who have had contact with hospital services within the past 12 months, and have one of the following diagnoses:
    - Dementia
    - Psychotic disorders
    - Mental disorders caused by drug misuse
    - Terminal cancer
- Estimated life expectancy less than a year based on prediction model
- Undergone major surgery in the past six months or have a planned surgery
- Severe hearing loss
- Language difficulties that require an interpreter
- Level of cognitive ability not sufficient for receiving and responding to telephone counselling
- No telephone connection
- Pregnancy

##### B.1.2 Randomisation

Patients who consented by returning their consent form were randomised. Patients were either allocated into the intervention group or into the control group using a 2:1 ratio, respectively, by using an online random sequence generator that was integrated into the electronic study management system. Patients randomised into the control group received a letter notifying them of this. Patients randomised into the intervention group were called to schedule a first meeting with a health coach. 2:1 randomisation was employed to increase patient recruitment and acceptability and on the recommendation of the ethics committee to maximise the number of patients receiving a potentially beneficial intervention they cannot access outside of the trial.

##### B.1.3 Sample size for RCT primary endpoint

Based on previous experience with studies among frequent emergency department visitors, the study is dimensioned to be able to detect what we deem is a clinically meaningful 12-15% reduction of the rate of occurrence of the primary outcome (i.e., emergency admissions) among the patients randomised to receive the intervention, compared to the control group. We anticipated a baseline rate of 1.5 emergency admissions per patient over the 2-year follow-up period and that the distribution of these events will approximately follow a Poisson distribution. Power calculations were conducted using simulations of 10,000 runs per simulation. We will include 2 participants per 1 control subject. In the targeted patient population, at a power of 0.90 and $\alpha$=0.048, this corresponds to a sample size of 1800 patients in the intervention arm and 900 controls for a total of 2700 patients. With further compensation for overdispersion, pertaining to the assumption of events following a Poisson distribution, and an expected

10% yearly drop-out, we estimated that we would need to randomise approximately 3000 patients. As suggested in main manuscript, reduced recruitment rates and positive outcomes led to early termination.

##### B.1.4 Patient recruitment and consent

All patients identified on the short list receive a generic invitation letter, a patient information sheet explaining the nature of the trial and health coaching intervention and a written consent form. Patients are contacted by a nurse trained HN employee within four days of the letter being sent to discuss the intervention further, confirm eligibility and suitability and obtain written consent for enrolment into the trial (See Figure B.2).

##### B.1.5 Intervention

The HN health coaches are all registered nurses or paramedics from a range of backgrounds including community and emergency care settings. They each undergo a bespoke accredited 6-month training course encompassing a range of well-defined theories covering motivational interviewing and personalised care plan development. They dedicate all their working time towards coaching patients in the trial. They do not have any role in primary care except that required by the study. The only collaboration that the coaches have with the patient's GP would be advising them of the patients' participation at the beginning of the intervention, advising that the patient had completed the intervention and any required contact to address specific patient issues e.g., acute deterioration.

Over the course of the trial, three health coaches and a site lead worked in the York team, where each coach followed up with the same patient over the whole study period. Each patient received an initial 1 – 1.5-hour face-to-face assessment meeting, followed by regular coaching calls over approximately a six-month timeframe at a frequency determined by the requirements of the individual patient. Coaches were instructed that the focus of the intervention was to provide empowerment, build confidence and improve health literacy to support patients to adopt a proactive approach to the self-management of their health.

The nature of the conversations focuses on identifying barriers to this approach, the cause of the health seeking behaviours and what their underpinning reasons are. Once the barriers and reasons were established, conversations would focus on addressing the barriers with the patient by way of goal setting, reducing inappropriate health seeking behaviours by improving the patients' health literacy, understanding how they could be pro-active in the self-management of their health, ensuring that they are on the correct care pathway, developing a sense of empowerment and building confidence. Motivational interviewing techniques are used to empower the patient and develop the patient's own motivation and willingness to engage in self-care. If the patient requires medical advice or treatment, then they are redirected to the GP or the hospital as appropriate.

#### B.2. Prediction model development and validation overview

A prediction model developed my Health Navigator (HN) was employed to identify patients at elevated risk of becoming high-intensity users of emergency and non-elective services. The prediction model was implemented via a software application that performs automated daily screening of hospital electronic health records (EHRs) and identifies high-risk patients.

Although the model development strategy was standardised across RCT sites (except mid-Essex where it was not feasible), each model was trained on 3 years of local hospital information. The training data was anonymised acute data, which focussed on the emergency department, in-patient and out-patient points of delivery. The model is fitted to the training data was a GLMnet (Lasso and elastic-net regularized linear model), and the hyperparameters were optimised using cross-validation to avoid statistical overfitting. GLMnet was shown to perform just as well as boosted approaches (e.g., XG Boost and Adaboost), but was chosen for its transparency and intelligibility. The model was internally validated using cross-validation and a split-sample approach. The training model achieved a discrimination performance of at least 0.8 on the test data across all sites included in the trial (excluding mid-Essex).

The prediction model itself uses a range of patient, in-patient and out-patient clinical information. More specifically, it utilises patient demographic information, emergency attendances and elective and non-elective admissions, outpatient visits, diagnostic codes, and discharge location. In addition, the model uses time-dependent variables to capture the change in hospital activity over time when assessing an individual's risk.

Regarding clinical diagnoses, HN's training data currently encompasses over 300 ICD-10 codes and 160 ECDS (SNOMED) codes to group patients' clinical conditions into 65 clinically meaningful groups (supported by medical professionals) across chronic, acute, and avoidable conditions.

To clarify the range of routinely collected patient information used in the prediction model, please find a list of common predictors included in the local prediction models.

1. Clinical conditions with which patients present:
    a. Cerebral ischaemia (stroke), hypertension, peripheral vascular disease
    b. Diabetes, cancer, asthma, chronic obstructive pulmonary disease
    c. Mental health (dementia, depression, schizophrenia) and learning disabilities (aphasia)
    d. Comorbidity severity defined by the Charlson comorbidity index
2. Count of emergency attendances and non-elective admissions in past 180/360 days
3. Means of attendance (e.g., emergency attendance via ambulance, did not attend outpatient appointments)
4. Demographics: ethnicity and age

### *Appendix C – Supplementary methods and findings*

#### C.1 Descriptive analysis

##### C.1.1 Methods

Supplementary Kaplan-Meier curves are provided in this section to illustrate crude differences in mortality rates across treatment groups, age, gender, and deprivation groups as discussed in the main manuscript.

##### C.1.2 Findings

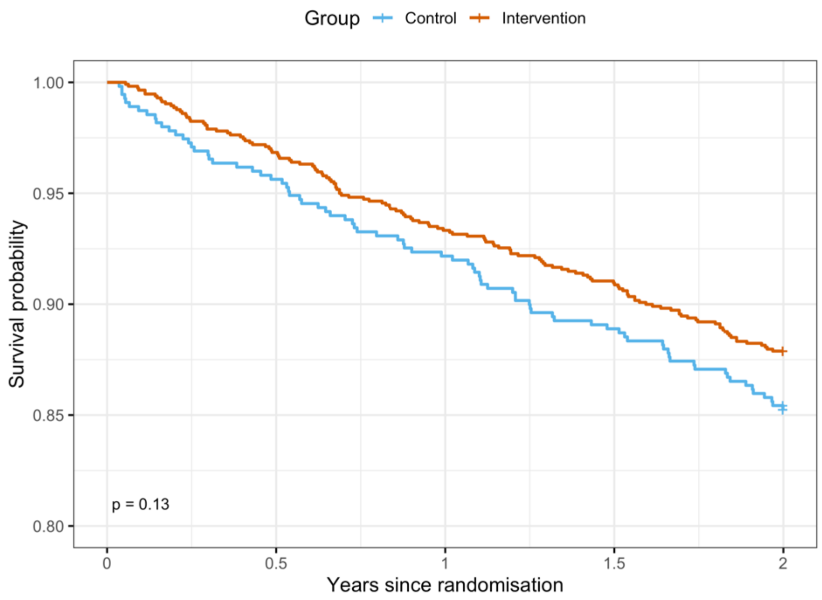

**Figure C.1: Kaplan-Meier survival curves by intervention group with corresponding p-value from a log-rank test.**

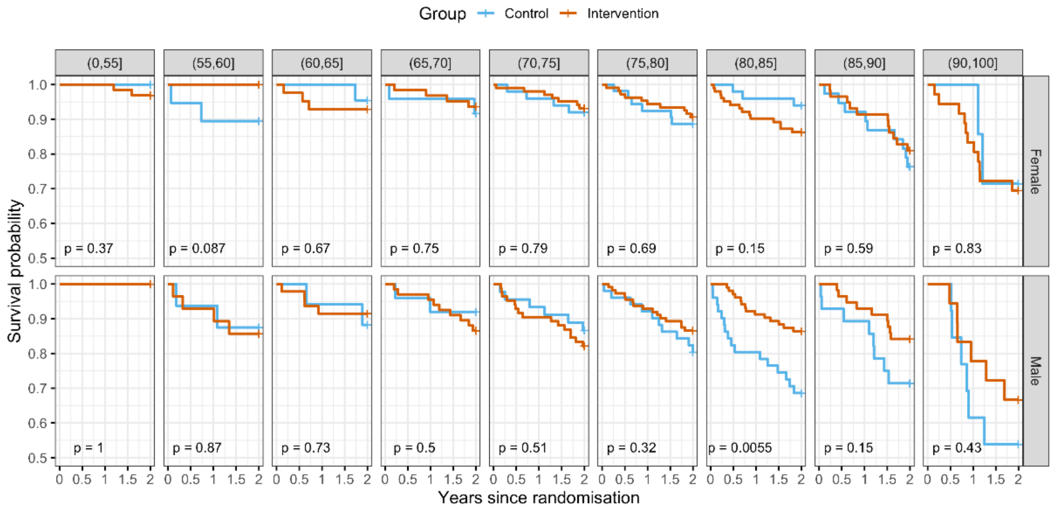

**Figure C.2: Kaplan-Meier survival curves by 5-year age band (panel columns) and gender (panel rows) with corresponding p-values from log-rank tests. Due to small sample sizes an underflow bin of 55 years and an overflow bin of 90 years were used.**

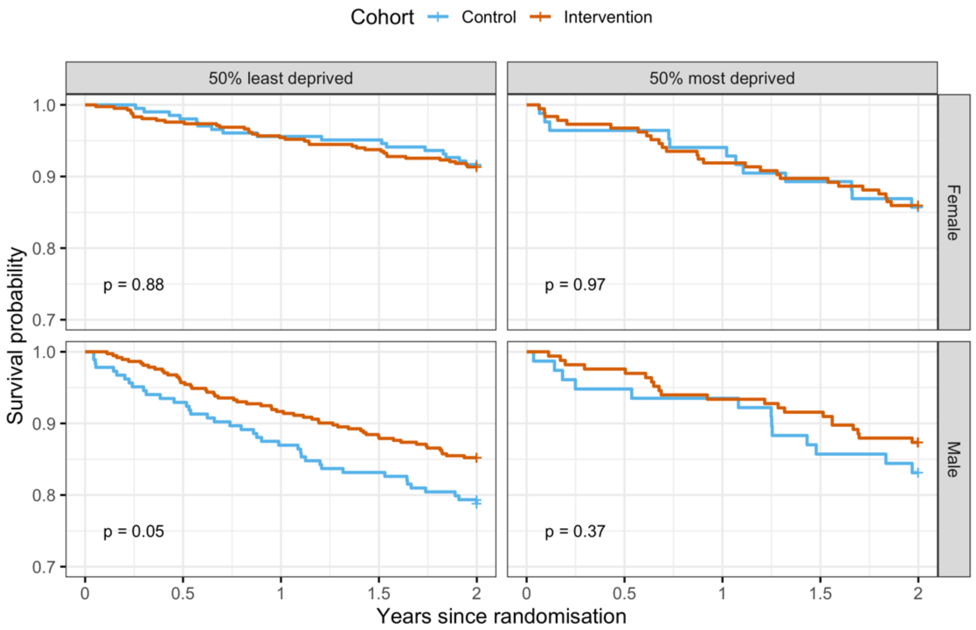

**Figure C.3: Kaplan-Meier survival curves by deprivation (panel columns) and gender (panel rows) with corresponding p-values from log-rank tests.**

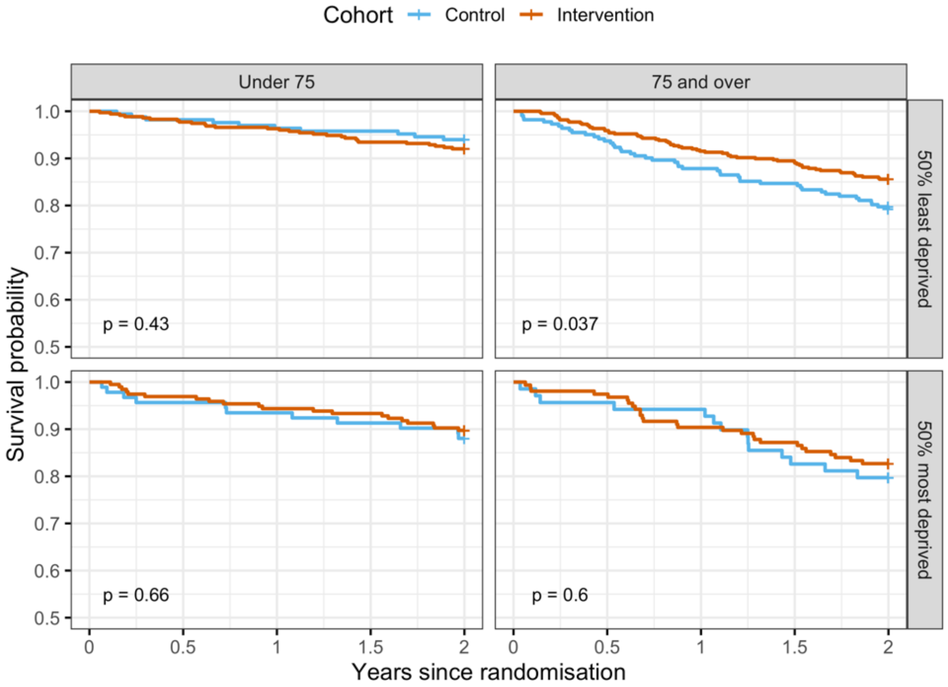

**Figure C.4: Kaplan-Meier survival curves by age band (panel columns) and deprivation (panel rows) with corresponding p-values from log-rank tests.**

#### C.2. Primary analysis

##### C.2.1 Methods

###### C.2.1.1 Informative prior derivation

As suggested in the main manuscript, when restricting the literature to existing interventions that are solely based in secondary care, their impact on mortality is limited. The primary outcomes of interest for these interventions are often patient-reported outcomes or healthcare utilisation [1-6]. Sometimes in-hospital mortality is included as a safety endpoint [7], or as a measure of health status to ensure matching across intervention and control subjects in observational studies [8-9].

The only studies identified to formally evaluate the impact of the intervention on mortality were a remote monitoring intervention for patients with multimorbidity [10], and a telephone-based intervention for older adults in managed care [11]. Another systematic review also included numerous interventions implemented in both primary and community care settings, where 21 out of the 36 studies included evaluated impact on patient mortality [12]. Sub-group analyses were also performed to explore whether elements of the study design or intervention impacted the conclusions from the meta-analyses performed.

As the evidence for mortality impact was limited, the informative prior for the intervention group in the Bayesian analysis were based on community-based interventions included in the systematic review [12], and the mortality impact evidence for the telephone-based intervention in older adults [11]. The remote monitoring intervention was too far removed from the intervention being evaluated in HN's randomised controlled trial [10]. Furthermore, as our study is also evaluating impact of the intervention on two-year mortality rates, this is referred to as 'long-term' mortality in the existing literature and will only be compared to existing studies alike [11, 12].

If we restricted the informative prior to be based on community-based interventions that targeted individuals that were identified by a prediction model and evaluated using a randomised controlled trial. The evidence base remained limited (see Table B.1).

| Trial | Year | Country | Effect type | Effect estimate | Confidence interval |
|---|---|---|---|---|---|
| Alkema et al. [11] | 2007 | USA | Odds Ratio (OR) | 0.55 | (0.36, 0.84) |
| Schraeder et al. [13] | 2001 | USA | Odds Ratio (OR) | 0.51 | (0.29, 0.91) |
| Toseland et al. [14] | 1996 | | Standardised mean difference | 0.19 | (-0.12, 0.5) |
| Stuck [15] | 2000 | Switzerland | Standardised mean difference | -0.26 | (-0.58, 0.06) |

**Table C.1 An overview of comparable case management studies and their effect estimates on long-term mortality (mortality over a duration longer than 12 months).**

For this reason, a wider perspective of the overall findings from the systematic review was employed to derive the informative prior of a null effect with a tighter confidence interval [12]. Thus, this included interventions implemented into primary care. More specifically, we focussed on the sub-group analysis presented in the 2015 meta-analysis and focussed on the standardised mean differences observed for those using a predictive algorithm (see Figures B.4 and B.5), and those evaluated via an RCT for long-term mortality.

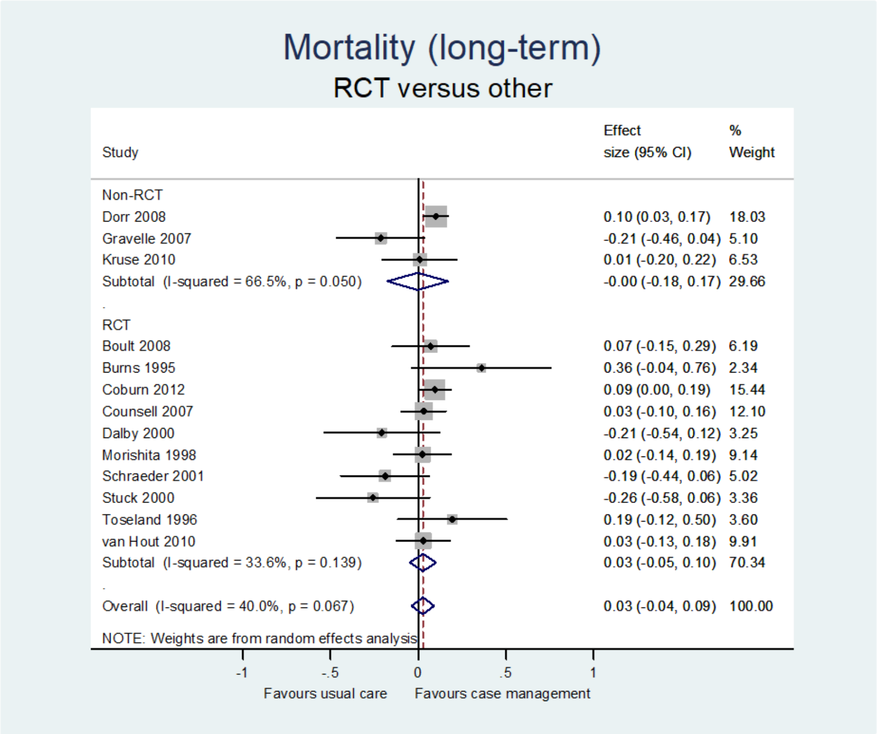

**Figure C.4 Sub-group analysis of intervention evaluation studies on mortality, evaluated by RCT and other means [12]. Figure presented in the supplementary findings of the cited journal article.**

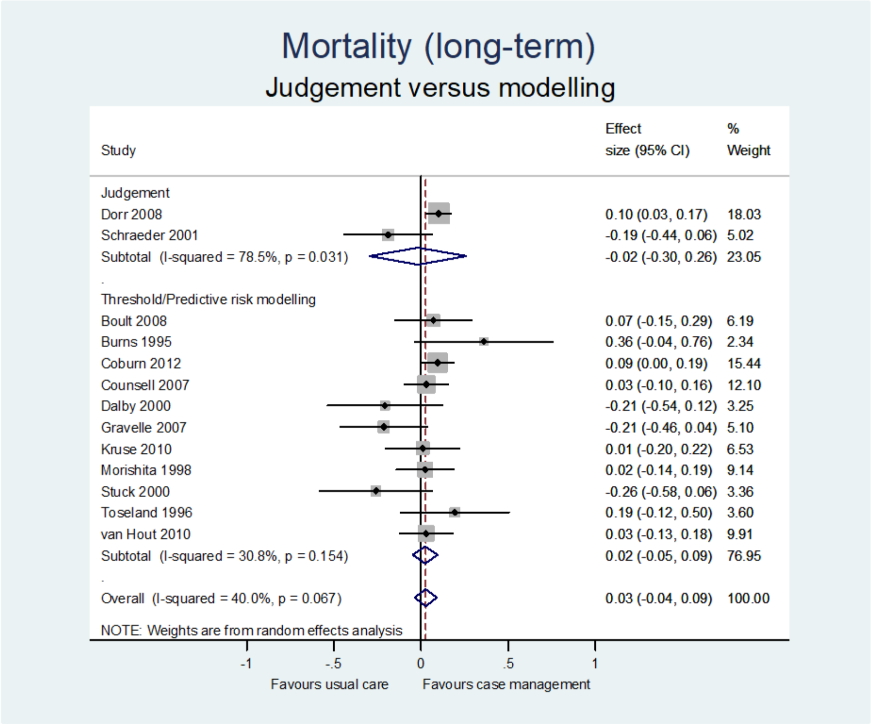

**Figure C.5 Sub-group analysis of intervention evaluation studies on mortality, with study subjects identified by a prediction model or otherwise [12]. Figure presented in the supplementary findings of the cited journal article.**

Utilising the information presented by Stokes et al., we hypothesised that the standardised mean difference in mortality between intervention and control groups would appear like the following:

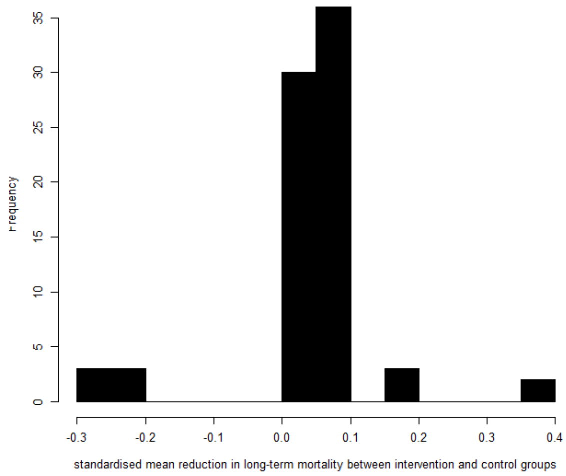

**Figure C.6 A histogram of standardised mean reductions in long-term mortality for interventions that targeted patients via prediction model and were evaluated via a randomised controlled study design.**

Utilising the existing evidence discussed in this sub-section, it was hypothesised that the intervention would have a null effect on mortality, and if it was to have a positive or negative effect, then it would not exceed extreme boundaries.

##### C.2.1.2 Bayesian Weibull Survival modelling

A Bayesian Weibull survival model was fitted with fixed effects for the intervention group (binary indicator), IMD decile less than or equal to 5 (binary indicator for living in the top 50% most deprived areas in the UK), age (in decades) and sex (binary indicator for males), and a random effect (frailty) for the intervention site. The random effect accounts for variation in the baseline hazard function between the different intervention sites and assumes proportionality between them. Non informative priors were placed on all fixed effects for control variables, random effects, and the Weibull shape parameter. An informative prior was placed on all fixed effects for the intervention and its interactions. The prior was chosen, based on previous studies and expert opinion, so that approximately 95% of the probability density function lies between hazard ratios 0.2 and 5. All parameters were estimated using the brms package in R [16].

The following Bayesian Weibull survival model was fitted to the data from all sites [17]:

If we denote, for patient $i$ , survival time as $t_i$, for the $n$ patients in the study population. The hazard function over time $t$ was modelled in the following way:

$$\lambda(t|\boldsymbol{x}_i) = \lambda_0(t\,|\boldsymbol{\theta})e^{\alpha_{site}+\,\beta_0\,group_i+\,\beta_1 sex_i+\beta_2 age_i+\,\beta_3 IMD_i} \quad (1)$$

Where, $\boldsymbol{\theta}$ is a vector of unknown parameters for the baseline hazard function and $\alpha_{site}$ denoted the frailty term specific to the hospital site of enrolment. $Group$, $age$, $sex$, and $IMD$ in **Equation 1** refer to the fixed effects, and the frailty term ($\alpha$) proportionally adjusts the parametric baseline hazard for each intervention site [18-19]. $\boldsymbol{x}_i$ denotes the vector of relevant (i.e., those defined in the exponent term) covariate observations for subject $i$.

For patient $i$, in **Equation 1,** $group$ refers to their allocated intervention group (1 = telephone-based case management intervention, 0 = control), $sex$ denotes whether the patient was male or not, $age$ was a continuous measurement of patient age in decades, $IMD$ denoted their 50% deprivation quantile (i.e., do they live in one of the top 50% most deprived areas?), and $site$ referred to the intervention site of they were enrolled onto the intervention.

For the Bayesian Weibull model, a general form of the Weibull baseline hazard function can be denoted as follows:

$$\lambda_0(t|\theta) = e^{\theta_1}t^{\theta_1 - 1}e^{\theta_2} \qquad (2)$$

Where $\theta = (\theta_1, \theta_2)$ are unknown parameters of the Weibull distribution that were estimated from the study data using Bayesian inference.

Interactions between the intervention and gender, age groups and deprivation bands were also investigated by adding in first order and second-order interactions to the above model. For first order and second order interactions, the exponent term in Equation 1 was adapted as suggested in Table B.2.

| Sub-groups of interest | Exponent term in Equation 1 |
|---|---|
| Gender | $\alpha_{site} + \boldsymbol{\beta_0}\, group_i * sex_i + \beta_1 sex_i + \beta_2 age_i + \beta_3 IMD_i$ |
| Age groups | $\alpha_{site} + \boldsymbol{\beta_0}\, group_i * age_categories_i + \beta_1 sex_i + \beta_2 age_i + \beta_3 IMD_i$ |
| IMD group | $\alpha_{site} + \boldsymbol{\beta_0}\, group_i * IMD_i + \beta_1 sex_i + \beta_2 age_i + \beta_3 IMD_i$ |
| Gender + Age groups | $\alpha_{site} + \boldsymbol{\beta_0}\, group_i * age_categories_i * sex_i + \beta_1 sex_i + \beta_2 age_i + \beta_3 IMD_i$ |
| Age groups + IMD group | $\alpha_{site} + \boldsymbol{\beta_0}\, group_i * age_categories_i * IMD_i + \beta_1 sex_i + \beta_2 age_i + \beta_3 IMD_i$ |
| Gender + IMD group | $\alpha_{site} + \boldsymbol{\beta_0}\, group_i * IMD_i * sex_i + \beta_1 sex_i + \beta_2 age_i + \beta_3 IMD_i$ |

**Table C.2 Exponent terms to clarify how pairwise interactions were included into the Bayesian Weibull Survival model ($age_categories$ refer to binary indicators for the following age bands: less than 65, 65 to < 75, 75 to < 85, 85 and over)**

Informative priors were employed for overall mortality impact. As there is no prior information specific to the subgroups investigated via pairwise interactions, the same informative prior was used as for the overall mortality impact.

The full specification of the Bayesian Weibull model used is as follows:

Linear predictor:

$$\mu_i = f(\alpha + \boldsymbol{\beta x_i} + \boldsymbol{u}\, \boldsymbol{z_i} + \epsilon_i),$$

where $\mu_i$ is the expected survival time for the $i^{th}$ patient, $\boldsymbol{\beta}$ and $\boldsymbol{u}$ are the fixed and random (site) effects respectively, $\boldsymbol{x_i}$ and $\boldsymbol{z_i}$ are the design vectors, $\epsilon_i$ is the random error, and f(.) is the link function:

$$f(.) = \frac{\exp(.)}{\Gamma\left(1 + \frac{1}{\sigma}\right)},$$

where $\Gamma$ is the gamma function and $\sigma$ is the Weibull shape parameter.

Model:

$$t_i|\sigma, \mu_i \sim Weibull(\sigma, \mu_i) \text{ for non-censored observations;}$$

$$p(t_i|\sigma, \mu_i) = WeibullCCDF(\sigma, \mu_i) \text{ for right-censored observations,}$$

where $WeibullCCDF$ is the Weibull complementary cumulative distribution function.

Priors:

$$p(\alpha) = StudentT(3, 0.7, 2.5);$$

$$p(\beta_i) = \begin{cases} Normal(0, 0.82) \text{ for intervention effects;} \\ Normal(0, 10) \text{ for control variables.} \end{cases}$$

$$p(u_i) = StudentT(3, 0, 2.5);$$

$$p(\epsilon_i) = StudentT(3, 0, 2.5);$$

$$p(\sigma) = Gamma(0.01, 0.01),$$

Where $t_i$ is the survival time of the $i^{th}$ patient.

Samples were drawn using Hamiltonian Monte Carlo (NUTS as implemented in Stan) with 8 chains, each with 2,500 warm-up iterations and 2,500 post warm-up iterations (20,000 total post warm-up draws). Other hyperparameters: adapt_delta=0.99, initial parameter values set to 0.

#### C.3 Sensitivity analysis

##### C.3.1 Methods

###### C.3.1.1 non-informative prior analysis

As discussed in Section B.1.1, prior evidence suggested a tighter confidence interval around a null effect of similar interventions on mortality. To investigate any bias introduced by the introduction of an informative prior, the following non-informative prior distribution was imposed for the hazard ratio of the intervention group on mortality:

$$\beta \sim N(0, 10)$$

Uniform non-informative priors were considered but it was unreasonable to apply an equal weighting to a hazard ratio of, for example, 1000 and zero, so a broad, normally distributed non-informative prior was chosen.

###### C.3.1.2 Cox proportional hazards analysis

Cox proportional hazards (PH) model was fitted with fixed effects for the intervention group (binary indicator), IMD decile less than or equal to 5 (binary indicator), age (in decades) and sex (binary indicator for males), and a random effect (frailty) for the intervention site. The random effect accounts for variation in the baseline hazard function between the different intervention sites and assumes proportionality between them.

The following Cox proportional hazards model was fitted to the data from all sites:

$$\lambda(t|\boldsymbol{x}_i) = \lambda_0(t)e^{\alpha_{site} + \beta_0 \, group_i + \beta_1 sex_i + \beta_2 age_i + \beta_3 IMD_i}$$

Where all parameters are defined in the same way as for the Bayesian model, except the baseline hazard ($\lambda_0(t)$) is not parametrically defined and is empirically defined using a step function of the event times in the observed data.

Pairwise interactions were explored for the Cox proportional hazards model in the same way they were explored using the Bayesian Weibull survival model.

C.3.2 Findings

| | **Hazard Ratio [95% CI]** | | |
|---|---|---|---|
| **Model 1 - Overall** | **Informative priors** | **Non-informative priors** | **Cox PH** |
| Intervention | 0.82 [0.62, 1.08] | 0.81 [0.61, 1.08] | 0.81 [0.62, 1.07] |
| Age (decades) | 1.60 [1.38, 1.89] | 1.60 [1.38, 1.89] | 1.58 [1.37, 1.83] |
| Gender=M | 1.66 [1.26, 2.23] | 1.66 [1.26, 2.21] | 1.64 [1.25, 2.15] |
| Deprivation=High | 1.24 [0.92, 1.67] | 1.24 [0.92, 1.67] | 1.24 [0.93, 1.66] |
| **Model 2 - By gender** | | | |
| Intervention:Male | 0.71 [0.50, 1.01] | 0.70 [0.48, 1.00] | 0.70 [0.49, 1.00] |
| Intervention:Female | 1.02 [0.67, 1.62] | 1.02 [0.65, 1.64] | 1.02 [0.65, 1.58] |
| Age (decades) | 1.60 [1.38, 1.90] | 1.60 [1.38, 1.89] | 1.58 [1.37, 1.83] |
| Gender=M | 2.10 [1.34, 3.44] | 2.12 [1.34, 3.49] | 2.08 [1.32, 3.28] |
| Deprivation=High | 1.25 [0.92, 1.69] | 1.25 [0.93, 1.69] | 1.25 [0.94, 1.68] |
| **Model 3 - By age categories** | | | |
| Intervention:Age[0,75) | 1.06 [0.70, 1.61] | 1.07 [0.69, 1.64] | 1.07 [0.70, 1.62] |
| Intervention:Age[75,100) | 0.72 [0.52, 0.99] | 0.71 [0.51, 0.99] | 0.72 [0.53, 0.98] |
| Age (decades) | 1.77 [1.46, 2.19] | 1.77 [1.46, 2.21] | 1.75 [1.45, 2.11] |
| Gender=M | 1.66 [1.27, 2.24] | 1.66 [1.27, 2.25] | 1.64 [1.26, 2.15] |
| Deprivation=High | 1.24 [0.91, 1.67] | 1.24 [0.92, 1.68] | 1.24 [0.93, 1.66] |
| **Model 4 - By deprivation bands** | | | |
| Intervention:HighDeprivation | 0.90 [0.55, 1.48] | 0.89 [0.54, 1.51] | 0.89 [0.54, 1.44] |
| Intervention:LowDeprivation | 0.79 [0.56, 1.10] | 0.78 [0.55, 1.10] | 0.78 [0.56, 1.09] |
| Age (decades) | 1.60 [1.38, 1.90] | 1.60 [1.38, 1.90] | 1.58 [1.37, 1.83] |
| Gender=M | 1.66 [1.26, 2.23] | 1.66 [1.26, 2.22] | 1.64 [1.25, 2.14] |
| Deprivation=High | 1.14 [0.69, 1.83] | 1.13 [0.69, 1.86] | 1.15 [0.71, 1.85] |
| **Model 4 - By age and gender** | | | |
| Intervention:Age[0,75):Female | 0.97 [0.52, 1.78] | 0.97 [0.48, 1.89] | 0.98 [0.51, 1.88] |
| Intervention:Age[75,100):Female | 1.03 [0.65, 1.65] | 1.03 [0.63, 1.70] | 1.02 [0.64, 1.65] |
| Intervention:Age[0,75):Male | 1.10 [0.68, 1.78] | 1.10 [0.66, 1.82] | 1.10 [0.67, 1.79] |
| Intervention:Age[75,100):Male | 0.57 [0.37, 0.84] | 0.54 [0.35, 0.83] | 0.55 [0.37, 0.83] |
| Age (decades) | 1.76 [1.45, 2.19] | 1.77 [1.45, 2.21] | 1.74 [1.44, 2.10] |
| Gender=M | 2.10 [1.36, 3.37] | 2.14 [1.35, 3.52] | 2.09 [1.32, 3.29] |
| Deprivation=High | 1.25 [0.92, 1.70] | 1.25 [0.93, 1.69] | 1.25 [0.93, 1.67] |

**Table C.3: An overview of findings using the Weibull Bayesian survival model (with informative and non-informative priors) and Cox PH model for both overall mortality and sub-group analyses**

| **Model** | **Hazard Ratio [95% CI]** |
|---|---|
| **Model 1 - Overall** | |
| Intervention | 0.74 [0.52, 1.05] |
| Age (decades) | 1.69 [1.39, 2.13] |
| Gender=M | 1.58 [1.12, 2.30] |
| Deprivation=High | 1.14 [0.76, 1.69] |
| **Model 2 - By gender** | |
| Intervention:Male | 0.60 [0.37, 0.94] |
| Intervention:Female | 1.02 [0.59, 1.81] |
| Age (decades) | 1.68 [1.39, 2.12] |
| Gender=M | 2.21 [1.25, 4.11] |
| Deprivation=High | 1.14 [0.77, 1.69] |
| **Model 3 - By age categories** | |
| Intervention:Age[0,75) | 1.04 [0.62, 1.74] |
| Intervention:Age[75,100) | 0.63 [0.41, 0.93] |
| Age (decades) | 1.93 [1.51, 2.59] |
| Gender=M | 1.59 [1.11, 2.33] |
| Deprivation=High | 1.14 [0.76, 1.70] |
| **Model 4 - By deprivation bands** | |
| Intervention:HighDeprivation | 0.78 [0.43, 1.49] |
| Intervention:LowDeprivation | 0.73 [0.48, 1.11] |
| Age (decades) | 1.69 [1.40, 2.13] |
| Gender=M | 1.58 [1.11, 2.32] |
| Deprivation=High | 1.09 [0.57, 2.00] |
| **Model 4 - By age and gender** | |
| Intervention:Age[0,75):Female | 1.00 [0.48, 2.07] |
| Intervention:Age[75,100):Female | 1.00 [0.56, 1.82] |
| Intervention:Age[0,75):Male | 1.04 [0.57, 1.88] |
| Intervention:Age[75,100):Male | 0.45 [0.26, 0.76] |
| Age (decades) | 1.90 [1.48, 2.53] |
| Gender=M | 2.16 [1.25, 3.90] |
| Deprivation=High | 1.13 [0.76, 1.68] |

| **Mortality rates and Numbers Needed to Treat** | | | |
|---|---|---|---|
| **Mortality deaths/1000/year (N)** | **Intervention** | **Control** | **2 year NNT** |
| Overall | 51.1 (872) | 69.3 (412) | 32 |
| Gender | | | |
| Female | 45.6 (458) | 44.7 (222) | * |
| Male | 57.4 (414) | 100.2 (190) | 14 |
| Age | | | |
| Under 75 | 38.2 (448) | 33.3 (203) | * |
| 75 and over | 65.1 (424) | 106.9 (209) | 15 |
| Deprivation | | | |
| 50% most deprived | 53.3 (276) | 69.3 (117) | 37 |
| 50% least deprived | 50.1 (596) | 69.4 (295) | 30 |
| Age & Gender | | | |
| Female, Under 75 | 24.6 (249) | 22.5 (114) | * |
| Female, 75 and over | 71.6 (209) | 68.9 (108) | * |
| Male, Under 75 | 55.7 (199) | 47.6 (89) | * |
| Male, 75 and over | 58.9 (215) | 151.8 (101) | 7 |
| * no reduction in mortality | | | |

**Table C.4: An overview of findings using the Bayesian Weibull survival model excluding Mid-Essex site for both overall mortality and sub-group analyses using an informative prior, as well as numbers needed to treat for sub-groups of interest.**

#### C.4 Secondary analysis to explain findings

##### C.4.1 Methods

###### C.4.1.1 Patient-reported living situation and medical conditions

For RCT participants in the intervention arm, the patient-reported living situation and clinical diagnoses were compared for those who were male aged 75 and over and the rest of the intervention arm. This data was collected from the patient during their clinical coaching intervention. Free text extraction of the ICD-10 codes was completed using the Medical Concept Annotation Tool (MedCAT), but the validity of SNOMED codes was not formally validated in our dataset [20].

###### C.4.1.2 Primary care consumption for a subset of York RCT participants

For a subset of RCT participants, who were based at the York site (for whom we had access to primary care records), crude incidence rates per patient year and incidence rate ratios, and corresponding 95% confidence intervals, for subgroups of interest (males and females under and over the age of 75, in both treatment arms) were tabulated and visualised using forest plots.

##### C.4.2 Findings

###### C.4.2.1 Patient-reported living situation and medical conditions

No noticeable differences between males aged 75 and over and the rest of the remaining intervention arm, regarding proportion of patients living alone and patient-reported health conditions.

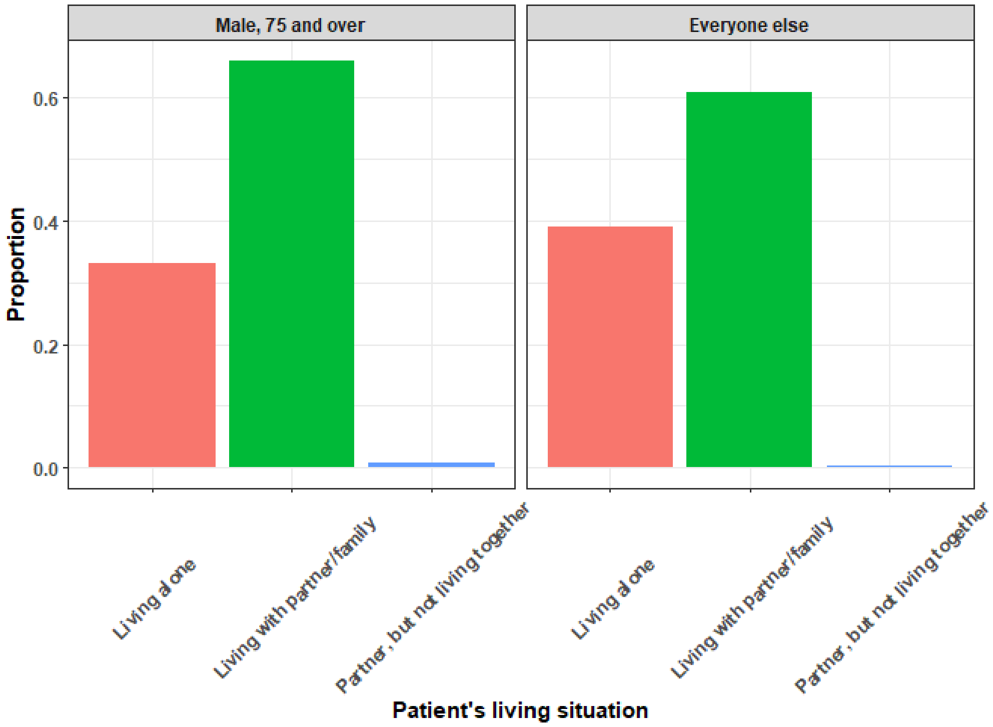

**Figure C.7: Comparing the proportion of subjects who are living alone in each of the sub-groups**

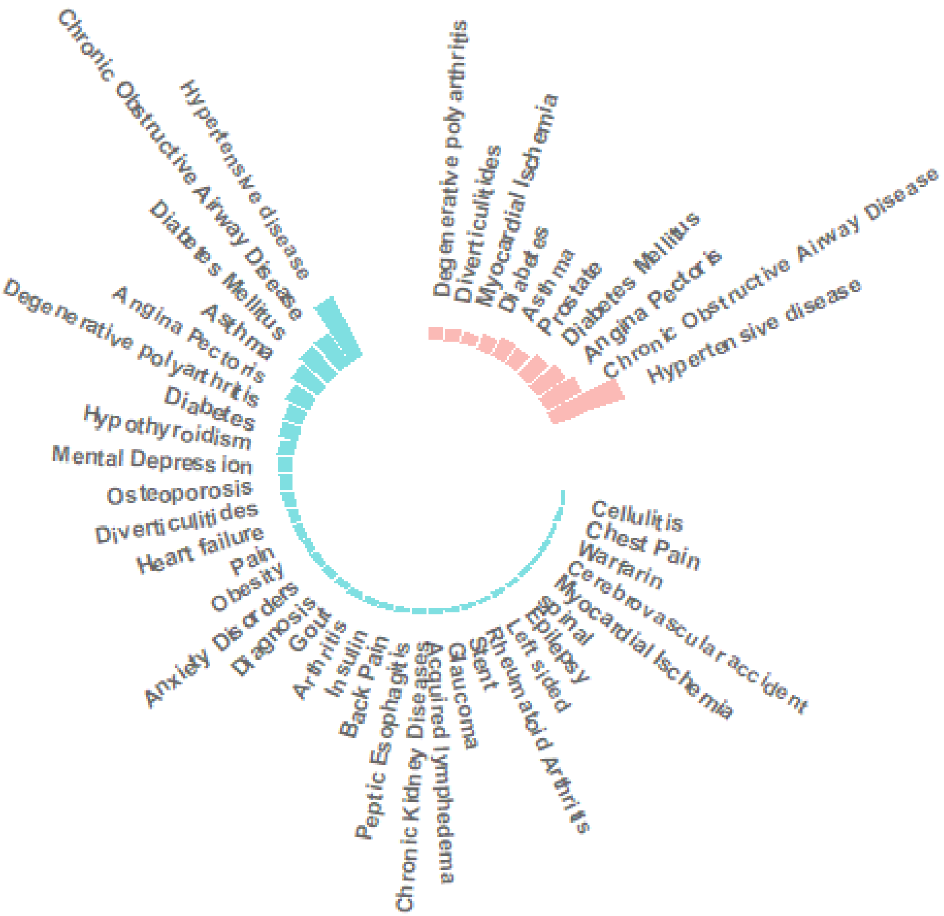

**Figure C.8 Comparing patient-reported current diseases across sub-groups (pink = male aged 75 and over, blue = rest of intervention arm)**

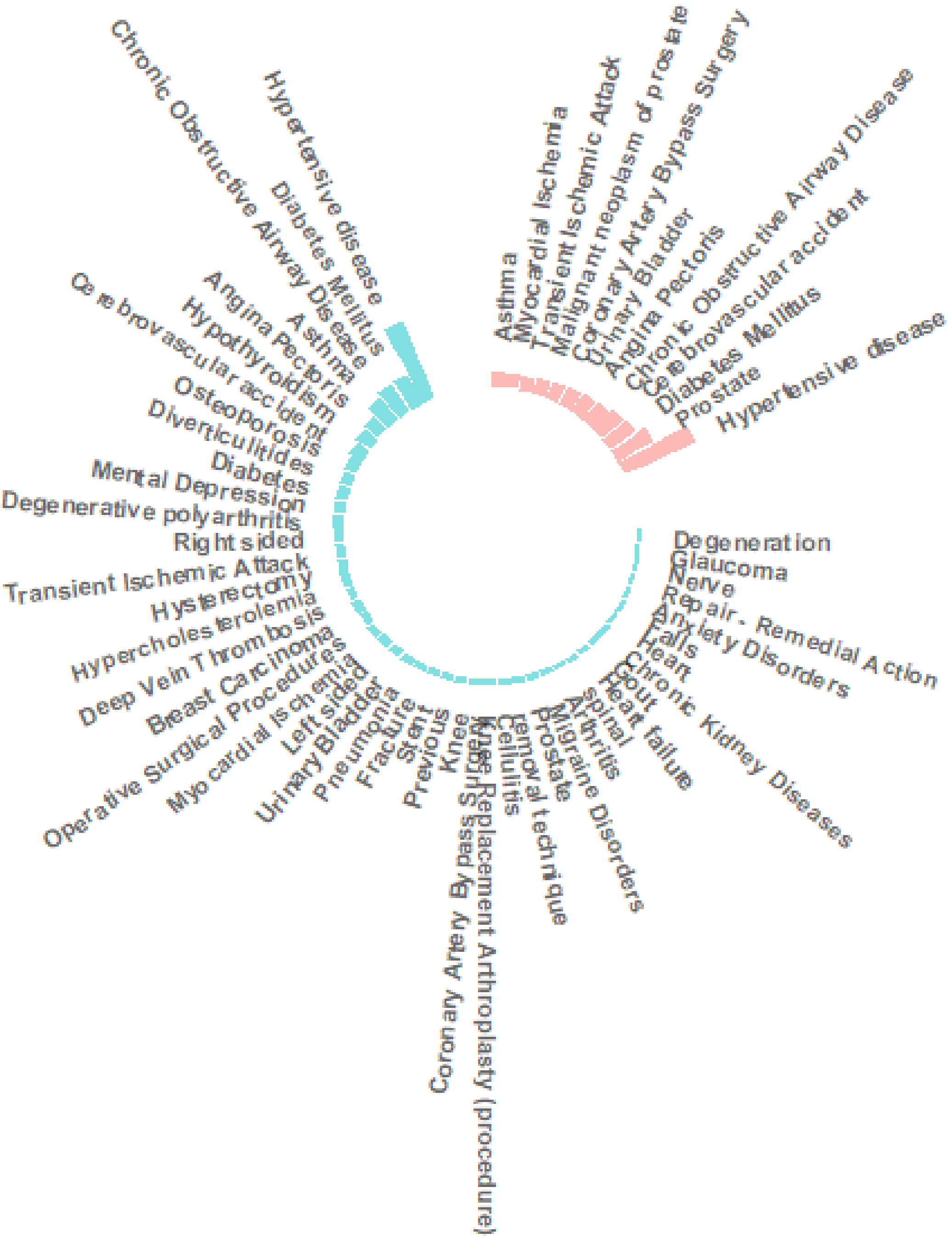

**Figure C.9 Comparing previous diseases across sub-groups (pink = male aged 75 and over, blue = rest of intervention arm)**

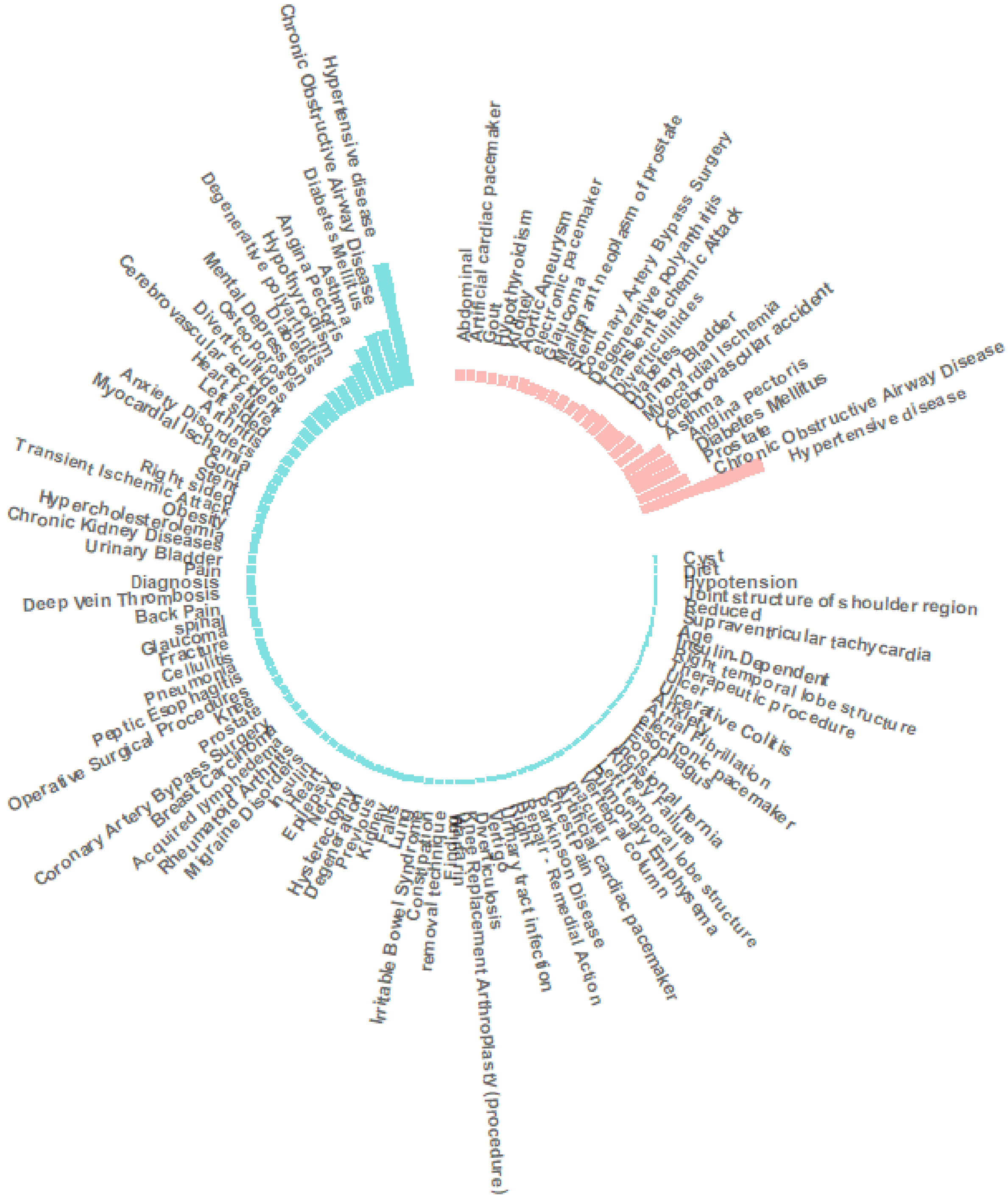

**Figure C.10: Comparing patient-reported current and past diseases across sub-groups of interest in the intervention arm (pink = male aged 75 and over, blue = rest of intervention arm)**

C.4.2.2 Primary care consumption for a subset of York RCT participants

Analysis is based on patients in the Vale of York RCT site, who we have access to their primary care records. The data on these patients has been sent from three different GP practices and contains 253 intervention patients and 110 control patients. Thus, this is only a subset of patients analysed in the mortality analysis, and only a subset of York patients.

Females aged 75 and over present the highest rate of primary care events compared to younger females and males who received the intervention, and control subjects who were also female and aged 75 and over. Males aged 75 and over present a significantly higher rate of primary care telephone interaction compared to the control arm, but this crude incidence rate is not observed to be higher than younger males or females in the intervention arm.

Caution: implicit assumptions about the mortality impact are being made here as the mortality data could not be linked. However, if there was evidence for the above hypothesis, we would observe a higher rate of primary care events for males aged over 75 compared to the remaining intervention subjects, and control patients.

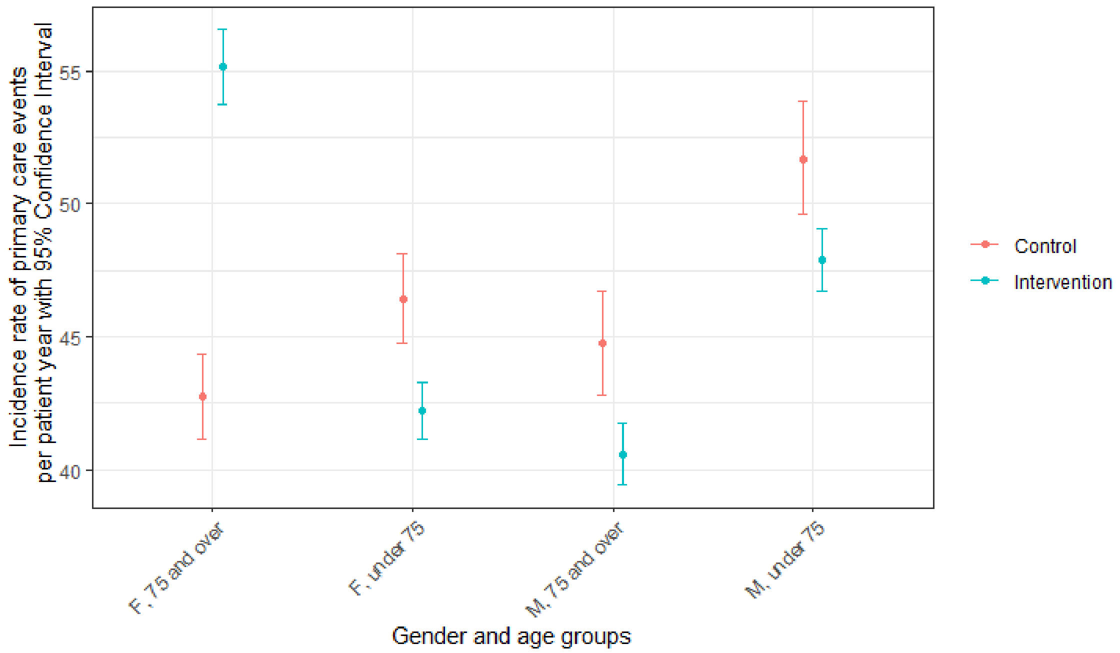

**Figure C.11: Incidence rates of all primary care events per patient year for intervention and control groups for the gender and age groups of interest, presented with their 95% confidence intervals.**

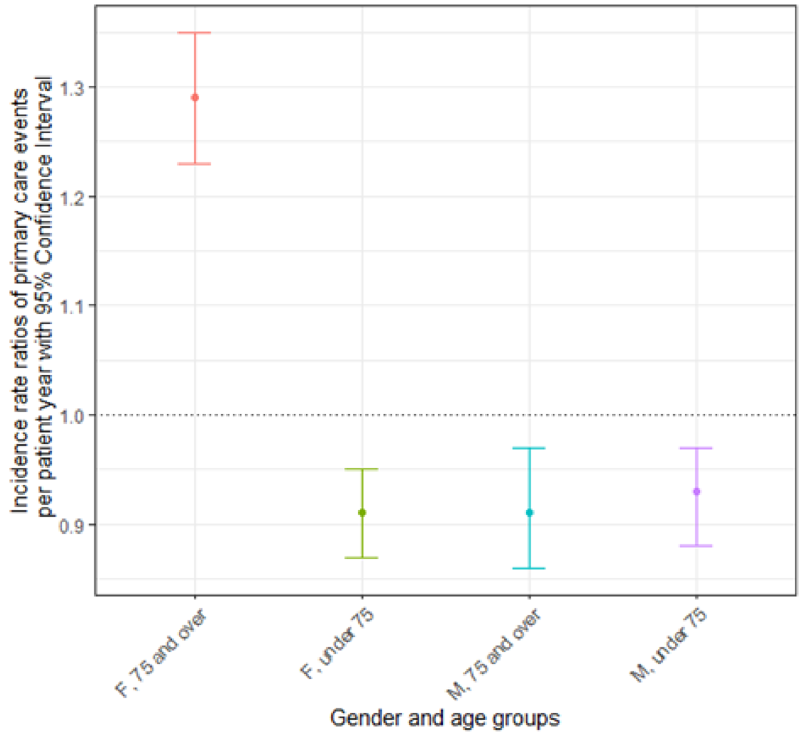

**Figure C.12: Incidence rate ratios of all primary care events per patient year for intervention and control groups for the gender and age groups of interest, presented with their 95% confidence intervals.**

| Subgroup | Incidence rate per patient year (95% confidence interval) | | Incidence Rate Ratio (95% confidence interval) |
|---|---|---|---|
| | Intervention | Control | |
| **All primary care events** | | | |
| Male, aged under 75 | 47.9 (46.7, 49.1) | 51.7 (49.6, 53.9) | **0.93 (0.88, 0.97)** |
| Male, aged 75 and over | 40.6 (39.5, 41.7) | 44.8 (42.8, 46.7) | **0.91 (0.86, 0.97)** |
| Female, aged under 75 | 42.2 (41.2, 43.3) | 46.4 (44.8, 48.1) | **0.91 (0.87, 0.95)** |
| Female, aged 75 and over | 55.1 (53.7, 56.5) | 42.8 (41.2, 44.4) | **1.29 (1.23, 1.35)** |
| **Face-to-Face** | | | |
| Male, aged under 75 | 24.3 (23.5, 25.2) | 26 (24.5, 27.6) | **0.93 (0.88, 1.0)** |
| Male, aged 75 and over | 23.5 (22.6, 24.3) | 28 (26.5, 29.6) | **0.84 (0.78, 0.9)** |
| Female, aged under 75 | 20.7 (19.9, 21.4) | 24.8 (23.6, 26.1) | **0.83 (0.78, 0.88)** |
| Female, aged 75 and over | 31.5 (30.4, 32.5) | 22.4 (21.2, 23.5) | **1.41 (1.32, 1.5)** |
| **Letter** | | | |
| Male, aged under 75 | 15.9 (15.2, 16.6) | 18.2 (17.0, 19.5) | **0.87 (0.80, 0.94)** |
| Male, aged 75 and over | 10.6 (10.0, 11.1) | 11.0 (10.0, 12.0) | **0.96 (0.87, 1.07)** |
| Female, aged under 75 | 13.6 (13.0, 14.2) | 12.6 (11.7, 13.5) | **1.08 (1.0, 1.1)** |
| Female, aged 75 and over | 13.9 (13.2, 14.6) | 11.1 (10.3, 12.0) | **1.25 (1.1, 1.4)** |
| **Telephone** | | | |
| Male, aged under 75 | 5.8 (5.4, 6.2) | 5.3 (4.6, 6.0) | **1.1 (0.95, 1.27)** |
| Male, aged 75 and over | 4.8 (4.5, 5.3) | 3.8 (3.3, 4.4) | **1.27 (1.07, 1.5)** |
| Female, aged under 75 | 5.9 (5.5, 6.3) | 6.7 (6.0, 7.3) | **0.89 (0.79, 1.27)** |
| Female, aged 75 and over | 7.7 (7.2, 8.2) | 7.1 (6.4, 7.7) | **1.09 (0.97, 1.22)** |
| **Referral** | | | |
| Male, aged under 75 | 1.9 (1.7, 2.2) | 2.2 (1.8, 2.7) | **0.87 (0.69, 1.10)** |
| Male, aged 75 and over | 1.7 (1.5, 1.9) | 1.9 (1.5, 2.4) | **0.88 (0.69, 1.14)** |
| Female, aged under 75 | 2.0 (1.8, 2.3) | 2.3 (2.0, 2.7) | **0.88 (0.72, 1.1)** |
| Female, aged 75 and over | 2.1 (1.8, 2.3) | 2.2 (1.8, 2.6) | **0.94 (0.76, 1.16)** |
| Female, aged 75 and over | 2.1 (1.8, 2.3) | 2.2 (1.8, 2.6) | **0.94 (0.76, 1.16)** |

**Table C.5: Incidence rates per patient year of all primary care events and sub-types of primary care events for males and females aged under and over 75 in the intervention and control groups, with the corresponding incidence rate ratios. [all metrics are reported with a 95% confidence interval)**

#### *Appendix D – CONSORT checklist*

Main manuscript pages and appropriate appendices in supplementary material provided.

| Section/Topic | Item No | Checklist item | Reported on page No |
|---|---|---|---|
| **Title and abstract** | | | |
| | 1a | Identification as a randomised trial in the title | 1 |
| | 1b | Structured summary of trial design, methods, results, and conclusions (for specific guidance see CONSORT for abstracts) | 2 |
| **Introduction** | | | |
| Background and objectives | 2a | Scientific background and explanation of rationale | 4 |
| | 2b | Specific objectives or hypotheses | 4 |
| **Methods** | | | |
| Trial design | 3a | Description of trial design (such as parallel, factorial) including allocation ratio | 4 |
| | 3b | Important changes to methods after trial commencement (such as eligibility criteria), with reasons | 4 |
| Participants | 4a | Eligibility criteria for participants | 4-5, Ap. B |
| | 4b | Settings and locations where the data were collected | 4, Ap. B |
| Interventions | 5 | The interventions for each group with sufficient details to allow replication, including how and when they were actually administered | 5, Ap. B |
| Outcomes | 6a | Completely defined pre-specified primary and secondary outcome measures, including how and when they were assessed | 5 |
| | 6b | Any changes to trial outcomes after the trial commenced, with reasons | 5 |
| Sample size | 7a | How sample size was determined | 5, Ap. B |
| | 7b | When applicable, explanation of any interim analyses and stopping guidelines | 5 |
| Randomisation: | | | 5, Ap. B |
| Sequence generation | 8a | Method used to generate the random allocation sequence | |
| | 8b | Type of randomisation; details of any restriction (such as blocking and block size) | 5, Ap. B |
| Allocation concealment mechanism | 9 | Mechanism used to implement the random allocation sequence (such as sequentially numbered containers), describing any steps taken to conceal the sequence until interventions were assigned | 5, Ap. B |
| Implementation | 10 | Who generated the random allocation sequence, who enrolled participants, and who assigned participants to interventions | 5, Ap. B |
| Blinding | 11a | If done, who was blinded after assignment to interventions (for example, participants, care providers, those assessing outcomes) and how | NA |
| | 11b | If relevant, description of the similarity of interventions | NA |
| Statistical methods | 12a | Statistical methods used to compare groups for primary and secondary outcomes | 6, Ap. C |
| | 12b | Methods for additional analyses, such as subgroup analyses and adjusted analyses | 6, Ap. C |

| | | | |
|---|---|---|---|
| **Results** | | | |
| Participant flow (a diagram is strongly recommended) | 13a | For each group, the numbers of participants who were randomly assigned, received intended treatment, and were analysed for the primary outcome | 7 |
| | 13b | For each group, losses and exclusions after randomisation, together with reasons | 7 |
| Recruitment | 14a | Dates defining the periods of recruitment and follow-up | 4 |
| | 14b | Why the trial ended or was stopped | 5 |
| Baseline data | 15 | A table showing baseline demographic and clinical characteristics for each group | 7 |
| Numbers analysed | 16 | For each group, number of participants (denominator) included in each analysis and whether the analysis was by original assigned groups | 7 |
| Outcomes and estimation | 17a | For each primary and secondary outcome, results for each group, and the estimated effect size and its precision (such as 95% confidence interval) | 9 |
| | 17b | For binary outcomes, presentation of both absolute and relative effect sizes is recommended | NA |
| Ancillary analyses | 18 | Results of any other analyses performed, including subgroup analyses and adjusted analyses, distinguishing pre-specified from exploratory | 10, Ap. C |
| Harms | 19 | All important harms or unintended effects in each group (for specific guidance see CONSORT for harms) | NA |
| **Discussion** | | | |
| Limitations | 20 | Trial limitations, addressing sources of potential bias, imprecision, and, if relevant, multiplicity of analyses | 11 |
| Generalisability | 21 | Generalisability (external validity, applicability) of the trial findings | 10 |
| Interpretation | 22 | Interpretation consistent with results, balancing benefits and harms, and considering other relevant evidence | 11 |
| **Other information** | | | |
| Registration | 23 | Registration number and name of trial registry | Integrated Research Application System project ID: 173319; and clinicaltrials.gov ID: 2015–000810-23 |
| Protocol | 24 | Where the full trial protocol can be accessed, if available | |
| Funding | 25 | Sources of funding and other support (such as supply of drugs), role of funders | 13 |
